# Sustainability and nutritional composition of food choices in hospital canteens: a pre-post intervention study

**DOI:** 10.64898/2026.04.02.26349952

**Authors:** Elisa Mansutti, Federica Fiori, Diana Menis, Peter Cautero, Caterina Liudmila Graziani, Daniela Zago, Marco Driutti, Lucia Lesa, Lucrezia Grillone, Francesca Cortelazzo, Angelica Cosolo, Manuela Mauro, Enrico Scarpis, Alessandro Conte, Maria Parpinel, Laura Brunelli

**Affiliations:** Department of Medicine, University of Udine, Udine, Italy; Health District of Gemona, Friuli Centrale Healthcare University Trust, Gemona, Italy; Medical Directorate, Hospital “Santa Maria della Misericordia” of Udine, Friuli Centrale Healthcare University Trust, Udine, Italy; Health District of Udine, Friuli Centrale Healthcare University Trust, Udine, Italy; Institute of Hygiene and Evaluative Epidemiology, Friuli Centrale Healthcare and University Trust, Udine, Italy; Food hygiene and nutrition Unit, Department of Prevention, Friuli Centrale Healthcare University Trust, Udine, Italy; Medical Directorate, Hospital of Latisana-Palmanova, Friuli Centrale Healthcare University Trust, Palmanova, Italy; Medical Directorate, Hospital of San Daniele-Tolmezzo, Friuli Centrale Healthcare University Trust, San Daniele, Italy; Accreditation, Quality, and Clinical Risk Unit, Friuli Centrale Healthcare University Trust, Udine, Italy

**Author notes:** these authors contributed equally (Elisa Mansutti, Federica Fiori). Corresponding author: Maria Parpinel, via Colugna 50, 33100 Udine, Italy. Mail.

**Keywords:** Food choices, Nutritional composition, Sustainability, Workplace canteen, Educational intervention, Environmental intervention

## Abstract

**Background:** Hospital canteens provide an effective setting to improve users’ dietary habits. The study evaluates users’ food choices – considering nutritional composition and environmental impact -after an educational and environmental intervention, comparing the results with pre-intervention choices.

**Methods:** A cross-sectional study was conducted in three hospital canteens (C1, C2, C3) in northeastern Italy, during two index weeks in September 2022 (T0) and 2023 (T1). An intervention was introduced between T0 and T1, consisting of posters on healthy eating, descriptive norm messages, and environmental changes regarding fruit and vegetables. Photos of lunch trays were collected, and choices were analyzed for nutritional composition and sustainability.

**Results:** 2,851 trays were analyzed: 1,227 at T0 (798 in C1, 228 in C2 and 201 in C3) and 1,624 at T1 (1,005 in C1, 348 in C2, 271 in C3). In C1 and C3, there was an increase in median energy (+30 kcal; +135 kcal) compared to pre-intervention meals, while in C2 there was a decrease (−118 kcal). Despite a slight improvement in macronutrient composition, at T1 meals in all canteens were still high in lipids (30%E; 39%E; 35%E) and low in carbohydrates (44%E; 39%E; 41%E). The fibre value fell within the recommended range only in C1 and C3. The median carbon (CF) and water (WF) footprints of meals in all canteens remained high: at T1 CF ranged from 966 gCO2eq. to 1,227 gCO2eq. and WF from 1,025 L H2O to 1,207 L H2O.

**Conclusion:** The intervention has led to partial improvements in food choices. To achieve more significant results, it may be necessary to implement a parallel intervention on food offer.

## 1 Introduction

Given the predicted 20% increase in demand for land-based animal products by 2050 (Food and Agriculture Organization of the United Nations 2023), the need to change eating habits is becoming increasingly urgent for both human and environmental health. The global aim is to gradually shift dietary habits, particularly by reducing red meat consumption, especially from ruminants, and encouraging the adoption of plant-based alternatives (Wynes and Nicholas 2017), which offer the greatest nutritional and environmental benefits (Project Drawdown 2020). In this context, public health policies that inform and guide consumers towards healthier and more sustainable diets are essential. While political action is crucial, it must be complemented by educational initiatives to inform people about the health and environmental effects of their diet. Consumers play a key role, as their choices significantly influence those of others, both through direct social interactions and via social networks (Althoff et al. 2017), and drive the market. In this context, canteens provide a particularly favorable environment for targeted interventions, as they are settings where an individual’s food choices can influence colleagues and the impact extends to many users. Consequently, an educational intervention in canteens may also have positive effects on those who have not directly benefitted from the intervention (Herman CP 2003). Nonetheless, changing deeply ingrained eating habits is one of the most complex challenges in public health. Several effective approaches in modifying food choices in the food service sector have been identified in the literature, including:

a. environmental changes: these involve altering the physical or social context independently of users, including modifications to the physical environment, infrastructure, processes, procedures, and social engagement (World Health Organization 2023). Examples include informative labelling of each dish, changes to food offerings (e.g., increasing the availability of healthy options), strategic food placement, pricing policies, and contextual regulations (e.g., prioritizing plant-based options in self-service). Environmental changes also include prompting, i.e., interventions in the physical environment (e.g., decorating spaces with references to plant-based foods) and enhancing the sensory and visual appeal of foods, including new preparation methods (Uglem et al. 2013; Anzman-Frasca et al. 2015; White et al. 2016);
b. knowledge and behavioral regulation: this includes interventions based on the dissemination of information through educational resources (posters/flyers, courses), the development of skills, and the use of incentives such as discounts, gadgets, or verbal prompts by canteen staff to encourage the desired behavior (Graça et al. 2023);
c. descriptive norm messages: messages based on social norms, as a nudging strategy, exploit the influence of the reference group to guide individual choices, correcting misperceptions about the behaviors of others (CialdiniR. B. and Trost M. R. 1998; Berkowitz 2005). Some authors suggest that descriptive norm messages can promote healthier and more sustainable food choices (Sharps and Robinson 2016; Thomas et al. 2017), although findings are still mixed, with some studies reporting limited or no significant effects (Papakonstantinou et al. 2025);
d. rewardings: these refer to the use of material (gadgets), financial, or verbal rewards following the achievement of specific behavioral goals (Graça et al. 2023).

Furthermore, food choices may also be indirectly modified by interventions in the food offer. Indeed, recent interventions in hospital cafeterias inspired by dietary models such as the *Planetary Health Diet* have demonstrated that increasing the availability of plant-based foods and reducing meat can improve the nutritional quality of meals, promote healthier choices, and reduce the environmental impact of food service (Li 2025). Hospital canteens or cafeterias, in particular, may serve as an optimal setting for the implementation of healthy and sustainable food policies that protect both human and environmental health, as hospitals are not only places of patient care but also major meal providers for employees. Studies by Jaworowska (Jaworowska et al. 2018) and Stern (Stern et al. 2021) highlight that food choices in hospitals are often nutritionally suboptimal and associated with a high environmental impact, indicating considerable scope for improvement. Similar evidence was also found in a previous work by the same research group as this study (Menis et al. 2024).

The aim of this study was to evaluate users’ choices regarding their nutritional composition and environmental impact of their meals after an educational and environmental intervention at the Friuli Centrale Healthcare University Trust (northeast Italy), and to compare the results with those from before the intervention.

## 2 Methods

This study is part of a larger project aimed at improving the nutritional and environmental awareness of hospital users, resulting from collaboration between the Department of Medicine at the University of Udine (Italy), catering service managers, the management of the Friuli Centrale Healthcare University Trust, and the local network of Health Promoting Hospitals and Health Services (HPH&HS). The results of the initial assessment of users’ food choices have already been published (Menis et al. 2024). This paper presents the results of a second assessment, conducted after an intervention aimed at promoting healthier and more sustainable food choices in the same hospital canteens. The methodology for data collection and analysis was consistent with that previously described (Menis et al. 2024).

### 2.1 Data collection

The study was conducted in three hospital canteens in the province of Udine: Udine (C1), Palmanova (C2), and San Daniele (C3). The canteens differ in terms of management - external for C1 and C2, internal for C3 - and daily attendance (approximately 450–480 users in C1, 100–120 in C2, and 90 in C3). The study population included hospital employees who used the canteen service on weekdays in September 2023, the same month investigated in the first survey (September 2022) to limit possible seasonal effects. Employees were allowed to select a combination of different courses for their meal according to Italian eating tradition, including a first course, second course, side dish (i.e. cooked vegetables) or salad, bread or substitutes, grated cheese, and dessert or fruit. As in the pre-intervention assessment, trays were marked with a unique code and photographed. During lunch, participants provided written informed consent and completed a questionnaire linked to the same code. The questionnaire collected socio-demographic and work-related data, as well as information on their perception of the intervention and their understanding of the messages conveyed. Participants whose questionnaires were returned empty were excluded from the analysis.

### 2.2 Intervention

The educational intervention comprised various elements, combined in a personalized manner for each canteen according to the specific organisation (type of catering management, internal vs external) and the structural possibilities of the spaces. For example, the recommended tray composition from a nutritional and sustainable perspective, created by the dietitians of the local Department of Prevention and presented as poster displayed in the canteen, was feasible only in the canteens with an external catering service and a rotational menu. In the internally managed canteen (C3), the menu was decided daily, making the recommended menu intervention inapplicable.

The elements of the intervention were as follows:

a. Educational element: different types of posters were displayed for two months before the week of analysis (T1) in the canteen spaces.
  i. Posters of the Mediterranean Diet Pyramid and Healthy Eating Plate: graphic representations of the Mediterranean Diet as a sustainable food model (Serra-Majem et al. 2020) and the Healthy Eating Plate created by the Department of Nutrition, Harvard T.H. Chan School of Public Health (Harvard T.H. Chan School of Public Health and Harvard Medical School). The number of posters varied for each canteen depending on the size of the room.
  ii. Posters with QR codes for further information on the food pyramid and ultraprocessed foods: these posters were in A4 format, one per canteen, displayed in a location easily visible to all canteen users.
  iii. Poster with the recommended composition of the tray (only possible in C1 and C2 as mentioned above): a daily poster with the recommended tray composition from a nutritional and sustainable perspective. The poster included the recommended composition for each day of the week. The posters were in A2 format and were changed weekly by the canteen staff.
b. Nudging with environmental restructuring element implemented about two months before T1 and included:
  i. Reversing the flow of dish choices: instead of starting with the choice of first and second courses, users were encouraged to begin with salad and side dishes before proceeding to other meal components. This intervention aimed to nudge the choice of vegetables over other food categories (Graça et al., 2023) and was introduced only in C2, where this structural change was possible.
  ii. Fruit, salad, and pulses self-service: in C1, salads, fruits, and pulses, instead of being served by canteen staff, were made available for self-service. Users could then decide their own portion. However, the limit of one fruit per user still applied.
c. Nudging with descriptive norm messages, based on the results of T0 and displayed as posters. For each canteen, the researchers calculated the percentages of users at T0 who chose fresh fruit as a dessert, who chose at least one portion of vegetables and, only for C1 where soft drinks were available, the percentage of users who chose water instead of soft drinks. One poster per descriptive norm message per canteen was displayed as close as possible to the relevant item. All descriptive norm message posters were displayed for about two months before the week of analysis. At the beginning of the survey week, these posters were removed to assess users’ knowledge gained after their exposure during the previous weeks.

In summary, the interventions for each canteen were as follows:

C1: educational posters; fruit, pulses, and salad self-service; posters with descriptive norm messages.

C2: educational posters; reversing the flow of dish choice; posters with descriptive norm messages.

C3: educational posters (except the suggested composition of the tray); posters with descriptive norm messages.

### 2.3 Statistical analysis

The recipes and standard portion sizes for each dish offered during the observation period were provided by the canteen staff. This information was matched at the ingredient level (Vásquez-Caicedo A. L. et al. 2008), with the foods in the Italian Food Composition Database for Epidemiological Studies (Gnagnarella P 2022) and of the SU-EATABLE LIFE dataset (Petersson et al. 2021) to obtain the energy and nutrient composition, as well as the environmental indicators (i.e., carbon footprint - CF; water footprint - WF) for each portion of each offered dish. Due to a lack of data in the SU-EATABLE LIFE dataset, we did not consider the post-marketing phases of each food’s life cycle in the environmental analysis, such as the cooking phase and (in C1 and C2) the transport from the cooking centre to the canteen, which occurred for some recipes. For the analysis of food choices, we considered all the dishes included in each tray.

As a second step, the analysis of the photos of the meal was performed. The researchers were trained to identify the standard portion for each food item by viewing a sample of photographs with different portion sizes. Users’ choices were visually estimated independently by two researchers to identify recipes and estimate portions compared to the standard portion (e.g., 50%, 100%, 150%). Discrepancies were resolved through discussion between the two researchers and a third researcher. By combining the data extracted from the photos with the recipe and food composition database, the researchers estimated the energy, nutritional profile, and environmental impact for each individual tray. Finally, the researchers compared the median values of CF and WF for each canteen with the reference ranges (CF: 800–1000 g CO_2_ equivalents/meal, WF: 700–1000 L H_2_0/meal) of the SU-EATABLE LIFE project (SU-EATABLE LIFE Project), while the median nutrient values were compared with the Italian DRVs (SINU Società Italiana di Nutrizione Umana 2014).

A descriptive analysis was conducted on participants’ characteristics associated with each tray, as well as users’ choices expressed in terms of energy and nutrient composition and the carbon and water footprint of the trays. Categorical variables were presented as absolute numbers and percentages. Nutritional and environmental variables were not normally distributed according to the Shapiro-Wilk test and were therefore reported as medians and 25th-75th percentiles. The Mann-Whitney U test was used to compare quantitative variables between T0 and T1 within each canteen. Differences in median values of nutritional and environmental variables across canteens were assessed using the Kruskal–Wallis test; when significant, post hoc comparisons were performed using Dunn’s test with Bonferroni adjustment. A p-value <0.05 was considered statistically significant. Tray categories (vegan, vegetarian, non-beef meat, fish, beef) were described using absolute and percentage frequencies. The trays were divided into the above categories as follows: “beef” if the tray contained at least one dish with beef; “fish” if the tray contained at least one fish dish; “non-beef meat” if there was at least one dish with meat excluding beef; “vegetarian” if there were no meat or fish dishes; “vegan” if there was no food source of animal origin on the tray. The chi-square test was used to assess associations between tray category and assessment time (T0 vs T1), and Pearson’s standardized residuals were examined. Similarly, the frequency of main-course choices within each category was calculated for each canteen, including a “no choice” option when no main course was selected. The chi-square test was also applied to evaluate associations between having a side dish and assessment time (T0 vs T1), and between having fruit and assessment time (T0 vs T1). Data were analysed with STATA (StataCorp. 2021. Stata Statistical Software: Release 17. College Station, TX: StataCorp LLC) and JASP Team (2024). JASP (Version 0.19.3) [Computer software].

All data were analyzed in aggregate form to ensure that the identity of individual participants could not be traced. The study was approved by the Institutional Review Board of the University of Udine (Italy) on 06/07/2022. All participants gave informed consent to take part in the study, and all procedures complied with the ethical standards set out in the Declaration of Helsinki.

## 3 Results

### 3.1 Characteristics of the study sample

A total of 1,624 lunch meals were analysed at T1 (1,005 from C1, 348 from C2, 271 from C3). In all canteens, participants were predominantly female (52% in C1, 58% in C2 and 63% in C3) and the mean age of participants was higher in C3 (48 ± 10 years) than in C2 (46 ± 11 years) and C1 (42 ± 12 years). Regarding professional profile, physicians was the most represented category in C1 (27%) and C3 (30%), while technician was the most represented in C2 (28%). The majority of participants were non-shift workers. A full description of the sample at T1, compared with those at T0, can be found in Table 1.

**Table 1.** Participant characteristics associated to each tray analyzed between T0 and T1, for each canteen.

|  | **C1** |  | **C2** |  | **C3** |  |
| --- | --- | --- | --- | --- | --- | --- |
|  | T0 (n = 798) | T1 (n = 1005) | T0 (n = 228) | T1 (n = 348) | T0 (n = 201) | T1 (n = 271) |
| **Age<sup>a</sup>** |  |  |  |  |  |  |
| ≤34 years | 239 (30) | 358 (35.6) | 71 (31.1) | 76 (21.8) | 26 (12.9) | 37 (13.7) |
| 35-54 years | 382 (47.9) | 467 (46.5) | 107 (46.9) | 183 (52.6) | 109 (54.9) | 153 (56.5) |
| ≥55 years | 177 (22.2) | 180 (17.9) | 50 (21.9) | 89 (25.6) | 66 (32.8) | 81 (29.9) |
| Missing | 0 (0) | 0 (0) | 0 (0) | 0 (0) | 0 (0) | 0 (0) |
| **Sex<sup>b</sup>** |  |  |  |  |  |  |
| Female | 402 (50.4) | 526 (52.3) | 150 (65.8) | 201 (57.8) | 140 (69.7) | 172 (63.5) |
| Male | 396 (49.6) | 479 (47.7) | 78 (34.2) | 147 (42.29) | 61 (30.4) | 99 (36.5) |
| Missing | 0 (0) | 0 (0) | 0 (0) | 0 (0) | 0 (0) | 0 (0) |
| **Professional profile<sup>b</sup>** |  |  |  |  |  |  |
| Physicians | 187 (23.4) | 268 (26.7) | 18 (7.9) | 32 (9.2) | 30 (14.9) | 82 (30.3) |
| Nurse | 96 (12) | 137 (13.6) | 62 (27.2) | 82 (23.6) | 47 (23.4) | 46 (17) |
| Health Care Assistant | 22 (2.8) | 63 (6.3) | 24 (10.5) | 24 (6.9) | 15 (7.5) | 25 (9.2) |
| Administrative staff | 174 (21.8) | 188 (18.7) | 44 (19.3) | 39 (11.2) | 43 (21.4) | 35 (12.9) |
| Technician | 209 (26.2) | 135 (13.4) | 46 (20.2) | 99 (28.5) | 21 (10.5) | 27 (10) |
| Others | 102 (12.8) | 206 (20.5) | 34 (14.9) | 72 (20.7) | 45 (22.4) | 47 (17.3) |
| Missing | 8 (1.0) | 8 (0.8) | 0 (0) | 0 (0) | 0 (0) | 9 (3.3) |
| **Type of work<sup>b</sup>** |  |  |  |  |  |  |
| Shift worker | 202 (25.3) | 285 (28.4) | 49 (21.5) | 88 (25.3) | 36 (17.9) | 85 (31.4) |
| Non-shift worker | 552 (69.2) | 660 (65.7) | 163 (71.5) | 255 (73.3) | 161 (80.1) | 164 (60.5) |
| Missing | 44 (5.5) | 60 (6) | 16 (7.0) | 5 (1.4) | 4 (2.0) | 22 (8.1) |

Notes: The values are presented as absolute frequencies and column percentages.
C1, canteen 1; C2, canteen 2; C3, canteen 3. \*The age of participants was normally distributed and therefore presented as mean and standard deviation.

Samples at T0 and T1 were comparable in terms of sex distribution across all canteens however, only in C3 there was no difference in age distribution, with the most represented age group being 35-54 years (54.2% at T0 and 56.5% at T1). In C1, although there was difference between the two assessments, the most represented age group was 35-54 years in both T0 and T1 (47.9% and 46.5% respectively), while the least represented was ≥55 years (22.2% and 17.9% respectively). In C2, the most represented age group was 35-54 years (46.9% and 52.6% respectively), while the least represented was ≥55 years at T0 (21.9%) and ≤34 years at T1 (21.8%). The type of work distribution was similar between T0 and T1 in C1 and C2; in C3, non-shift workers increased by 13.5%. Professional profile distribution was different between the two assessments in all three canteens.

### 3.2 Nutritional values and sustainability of food choices in the post-intervention phase

The energy, nutrient content and the environmental impact of the trays at T1 differed between the three canteens (p < 0.05), except for saturated fatty acids (SFA) (see Table 2). In particular, the median energy content of the trays was higher in C1 (884 kcal/tray) compared to C2 (782 kcal/tray) and C3 (781 kcal/tray). This difference was significant in the Dunn test between C1 and C2, and between C1 and C3 (Supplementary Table 1). Most differences in macronutrients were observed between C1 and C2 (p <0.001), except for SFA and monounsaturated fatty acids (MUFA) content. The highest carbohydrate and protein contents were found in C1 (carbohydrates: 104.4 g/tray, 45%E; proteins: 43.2 g/tray, 20%E), while the highest lipid content was found in C2 (33.2 g/tray, 39%E).

**Table 2.** Energy, nutrients content and sustainability indicators of the choices in C1, C2 and C3 at T1 expressed as medians, 25^th^ and 75^th^ percentiles.

| Variables | C1 (n = 1005) | C2 (n = 348) | C3 (n = 271) | p value<sup>a</sup> |
| --- | --- | --- | --- | --- |
| **Energy (kcal/tray)** | 884 [705–1083] | 782 [624–925] | 781 [615–935] | 0.0001 |
| **Protein (%E)** | 20 [15–25] | 19 [15–23] | 20 [17–24] | 0.0189 |
| **Lipids (%E)** | 30 [25–38] | 39 [34–46] | 35 [31–41] | 0.0001 |
| SFA (%E) | 7 [5–9] | 8 [7–10] | 9 [6–10] | 0.0001 |
| MUFA (%E) | 15 [12–19] | 18 [15–23] | 19 [16–23] | 0.0001 |
| PUFA (%E) | 5 [3–6] | 7 [5–9] | 5 [3–8] | 0.0001 |
| **Carbohydrates (%E)** | 45 [36–53] | 39 [32–46] | 41 [34–46] | 0.0001 |
| Sugars (%E) | 11 [8–16] | 9 [6–13] | 11 [9–15] | 0.0001 |
| **Fibre (%E)** | 3 [2–3] | 2 [2–3] | 3 [2–4] | 0.0001 |
| **Protein (g)** | 43.2 [31.4–57.3] | 34.5 [28.1–44.4] | 38.2 [29.4–49.1] | 0.0001 |
| **Lipids (g)** | 28.9 [22.2–38.9] | 33.2 [24.8–42.9] | 30.4 [23.9–37.5] | 0.0001 |
| SFA (g) | 6.5 [4.6–10.0] | 6.7 [4.8–9.9] | 7.1 [4.8–10.1] | 0.6026 |
| MUFA (g) | 14.7 [11.5–18.6] | 15.0 [11.4–19.9] | 16.0 [12.6–20.7] | 0.0120 |
| PUFA (g) | 4.5 [3.1–6.3] | 5.7 [4.2–8.1] | 3.7 [2.9–6.8] | 0.0001 |
| EPA + DHA (mg) | 58 [0–252] | 25 [0–116] | 30 [0–95] | 0.0001 |
| **Carbohydrates (g)** | 104.4 [76.3–141.7] | 79.7 [58.5–103.0] | 80.7 [64.1–105.6] | 0.0001 |
| Sugars (g) | 26.7 [17.8–38.8] | 18.3 [11.2–26.3] | 24.0 [18.8–30.2] | 0.0001 |
| **Fibre (g)** | 11.1 [7.6–15.0] | 7.8 [5.5–11.7] | 11.3 [8.6–15.3] | 0.0001 |
| **CF (g CO<sub>2</sub>eq./tray)** | 1227 [897–1756] | 966 [638–1284] | 1126 [775–1585] | 0.0001 |
| **WF (L H<sub>2</sub>O/tray)** | 1207 [952–1557] | 1025 [767–1311] | 1195 [881–1641] | 0.0001 |

Notes:<sup>a</sup> = Kruskal-Wallis test

From a sustainability perspective (Table 2), the median values of CF and WF were higher in C1 (CF 1227 g CO_2_eq./tray; WF 1207 L H_2_O/tray) than in C3 (CF 1126 g CO_2_eq./tray; WF 1195 L H_2_O/tray) and C2 (CF 966 g CO_2_eq./tray; WF 1025 L H_2_O/tray). This difference was statistically significant (p < 0.026) between all canteens (Table 3).

**Table 3.** Trays containing fruit, side dish and salad at T0 and T1, reported as absolute and relative frequencies.

| Fruit | C1 |  |  |  | C2 |  |  |  | C3 |  |  |  |
| --- | --- | --- | --- | --- | --- | --- | --- | --- | --- | --- | --- | --- |
|  | T0 | T1 | Total | p-value<sup>b</sup> | T0 | T1 | Total | p-value<sup>b</sup> | T0 | T1 | Total | p-value<sup>b</sup> |
|  |  |  |  | p = 0.062 |  |  |  | p = 0.855 |  |  |  | p = 0.644 |
| Yes | 456 (57.1) | 530 (52.7) | 986 |  | 102 (44.7) | 153 (44) | 255 |  | 155 (77.1) | 204 (75.3) | 359 |  |
| No | 342 (42.9) | 475 (47.3) | 817 |  | 126 (55.3) | 195 (56) | 321 |  | 46 (22.9) | 67 (24.7) | 113 |  |
| Total (n) | 798 | 1005 | 1803 |  | 228 | 348 | 576 |  | 201 | 271 | 472 |  |
| Side dish | T0 | T1 | Total | p-value<sup>b</sup> | T0 | T1 | Total | p-value<sup>b</sup> | T0 | T1 | Total | p-value<sup>b</sup> |
|  |  |  |  | p = 0.020 |  |  |  | p = 0.980 |  |  |  | p = 0.011 |
| Yes | 531 (66.5) | 720 (71.6) | 1251 |  | 128 (56.1) | 195 (56) | 323 |  | 128 (63.7) | 202 (74.5) | 330 |  |
| No | 267 (25.6) | 285 (28.4) | 552 |  | 100 (43.9) | 153 (44) | 253 |  | 73 (36.3) | 69 (25.5) | 142 |  |
| Total (n) | 798 | 1005 | 1803 |  | 228 | 348 | 576 |  | 201 | 271 | 472 |  |
| Salad | T0 | T1 | Total | p-value<sup>b</sup> | T0 | T1 | Total | p-value<sup>b</sup> | T0 | T1 | Total | p-value<sup>b</sup> |
|  |  |  |  | p < 0.001 |  |  |  | p = 0.049 |  |  |  | p = 0.034 |
| Yes | 186 (23.3) | 401 (39.9) | 587 |  | 116 (50.9) | 206 (59.2) | 322 |  | 66 (32.8) | 115 (42.4) | 181 |  |
| No | 612 (76.7) | 604 (60.1) | 1216 |  | 112 (49.1) | 142 (40.8) | 254 |  | 135 (67.2) | 156 (57.6) | 291 |  |
| Total (n) | 798 | 1005 | 1803 |  | 228 | 348 | 576 |  | 201 | 271 | 472 |  |

Notes: The values are presented as absolute frequencies and column percentages; <sup>b</sup> Chi-squared test

As the intervention was implemented in different combinations across the three canteens, and the canteens themselves are not comparable in terms of food offer, management type, and user rates, we analyzed each canteen separately.

Figure 1 shows that in C1, the median energy intake of the meal increased in the post-intervention measurement compared to T0 (854 kcal at T0 vs. 884 kcal at T1; p < 0.001). Considering the content in grams per tray, carbohydrates (103 g at T0 vs. 104.4 g at T1; p < 0.002) and fibre (9.9 g at T0 vs. 11.1 g at T1; p < 0.001) increased (Supplementary Table 2), while protein (45.7 g at T0 vs. 43.2 g at T1; p = 0.083) and lipids (28.9 g at T0 vs. 29.9 g at T1; p = 0.743) remained unchanged. As the energy content of the meal increased, the energy contribution of carbohydrates did not increase significantly (44%E at T0 vs. 45%E at T1; p = 0.639), while the energy from protein (21%E at T0 vs. 20%E at T1; p < 0.001) and lipids (32%E at T0 vs. 30%E at T1; p = 0.002) decreased. Similarly, although the content of both SFA and MUFA in grams increased in the meal at T1, only the energy contribution of MUFA increased, while that of SFA decreased (p < 0.001). Regarding sustainability, CF decreased significantly between T0 and T1 (1338 g CO_2_ eq./meal at T0 vs. 1227 g CO_2_ eq./meal at T1; p < 0.001), despite the total energy of the meal increased (Figure 1). The change in WF, however, was not significant (p = 0.107).

**Fig. 1.**
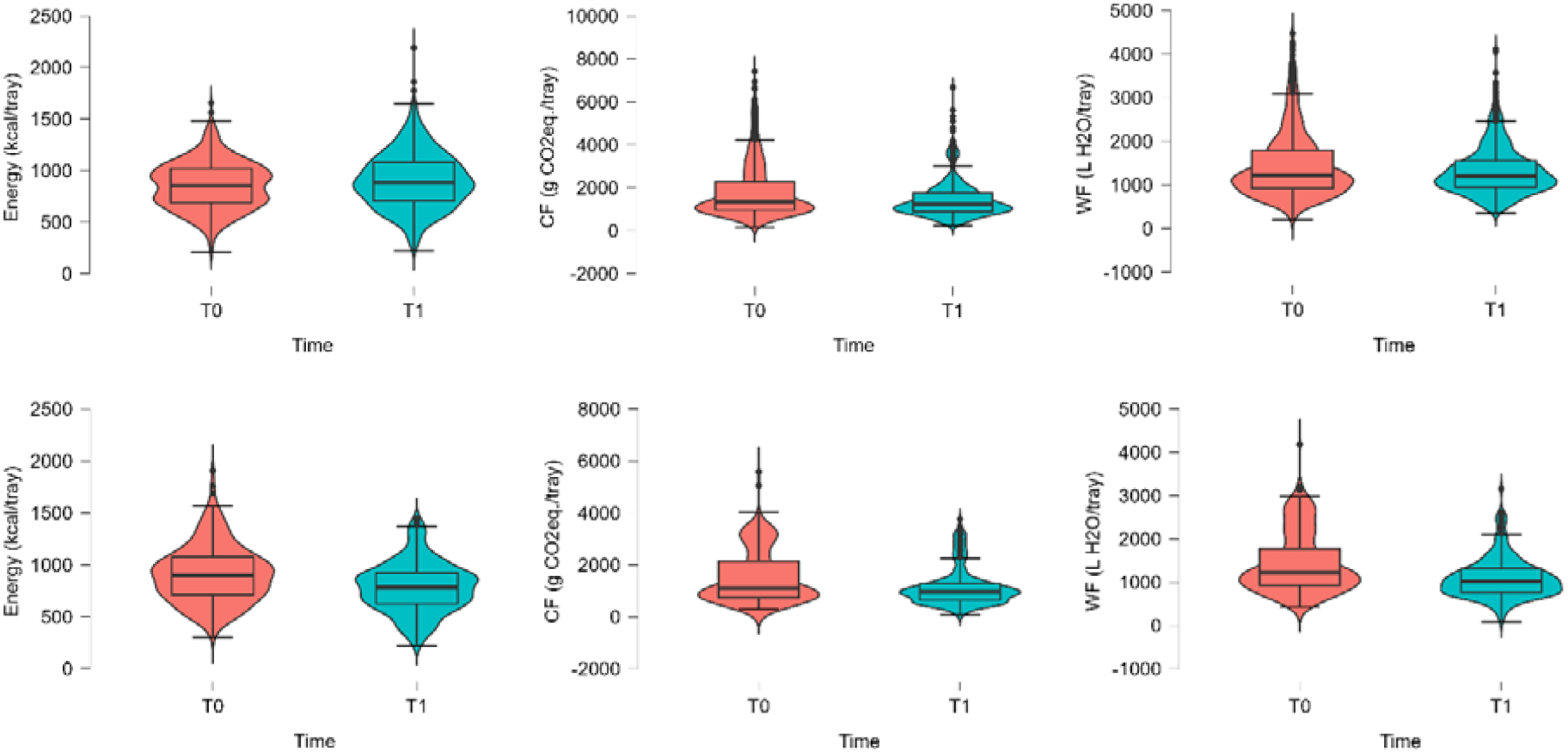
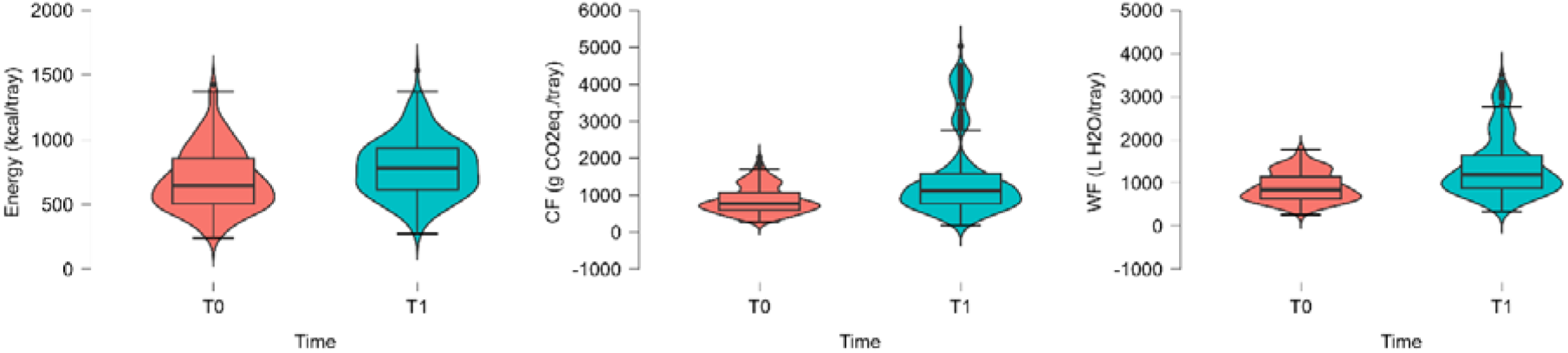
Distribution of energy values, CF and WF for C1 (the first row), C2 (the second row) and C3 (the third row) at T0 and T1

In C2, the median energy intake decreased (900 kcal at T0 vs. 782 kcal at T1; p < 0.001), accompanied by a decrease in grams of carbohydrates (93.1 g at T0 vs. 79.7 g at T1; p < 0.001), protein (37.5 g at T0 vs. 34.5 g at T1; p = 0.030), lipids (37.7 g at T0 vs. 33.2 g at T1; p < 0.001), and fibre (10.1 g at T0 vs. 7.8 g at T1; p < 0.001). Only long-chain omega-3 fatty acids (EPA+DHA) increased (p < 0.021). In terms of energy contribution, carbohydrates remained unchanged (39%E at T0 vs. 39%E at T1; p = 0.265), as did lipids (40%E at T0 vs. 39%E at T1; p = 0.806), while the contribution of protein increased (17%E at T0 vs. 19%E at T1; p < 0.001). There was a decrease in the contribution of SFAs (p = 0.009), an increase in that of polyunsaturated fatty acids (PUFAs) (p < 0.001), while MUFAs remained unchanged (p = 0.067). In parallel to the decrease in meal energy, both CF and WF decreased (CF: 1094 g CO_2_ eq./meal at T0 vs. 966 g CO_2_ eq./meal at T1; WF: 1220 at T0 vs. 1025 at T1; p < 0.001).

In C3, energy intake increased (646 kcal at T0 vs. 781 kcal at T1; p < 0.001). There was an increase in grams of protein (32.4 g at T0 vs. 38.2 g at T1; p < 0.001), total lipids (25.4 g at T0 vs. 30.4 g at T1; p < 0.001), MUFA and PUFA, and fibre (8.5 g at T0 vs. 11.3 g at T1; p < 0.001). There was also an increase in the energy contribution of lipids (34%E at T0 vs. 35%E at T1; p = 0.004) and a decrease in the contribution of carbohydrates (44%E at T0 vs. 41%E at T1; p < 0.001). For the energy contribution of fatty acids, there was a decrease in SFA (p = 0.015) and an increase in MUFA and PUFA (p < 0.001 and p = 0.016, respectively). CF and WF both increased (CF: 773 g CO_2_ eq./meal at T0 vs. 1126 g CO_2_ eq./meal at T1; WF: 847 at T0 vs. 1195 at T1; p < 0.001). The comparison of nutritional values and sustainability of food choices between T0 and T1 in the three canteens (C1, C2, and C3) can be found in Supplementary Table 2.

The full charts showing the distribution of nutritional and sustainability values are available in Supplementary Figures 1, 2 and 3.

### 3.3 Food choices

Figure 2 shows the categorization of trays in the pre- and post-intervention phases. A significant association between the distribution of tray categories and assessment period (T0-T1) was found for all three canteens (p < 0.001). In C1, during the post-intervention phase, trays categorized as fish increased (14.5% at T0 vs 36.5% at T1), while those categorized as meat without beef (38.7% at T0 vs. 33.9% at T1) and beef (31.5% at T0 vs. 11.4% at T1) decreased. In C2, there was a significant increase in trays categorized as meat without beef (17.1% at T0 vs. 31.6% at T1) and a decrease in those with beef (28.8% at T0 vs. 15.5% at T1). In C3, there was a significant decrease in fish trays (70.2% at T0 vs. 37.6% at T1) and an increase in beef trays (0% at T0 vs. 28.4% at T1). The complete results are available in Supplementary Table 3.

**Fig. 2.**
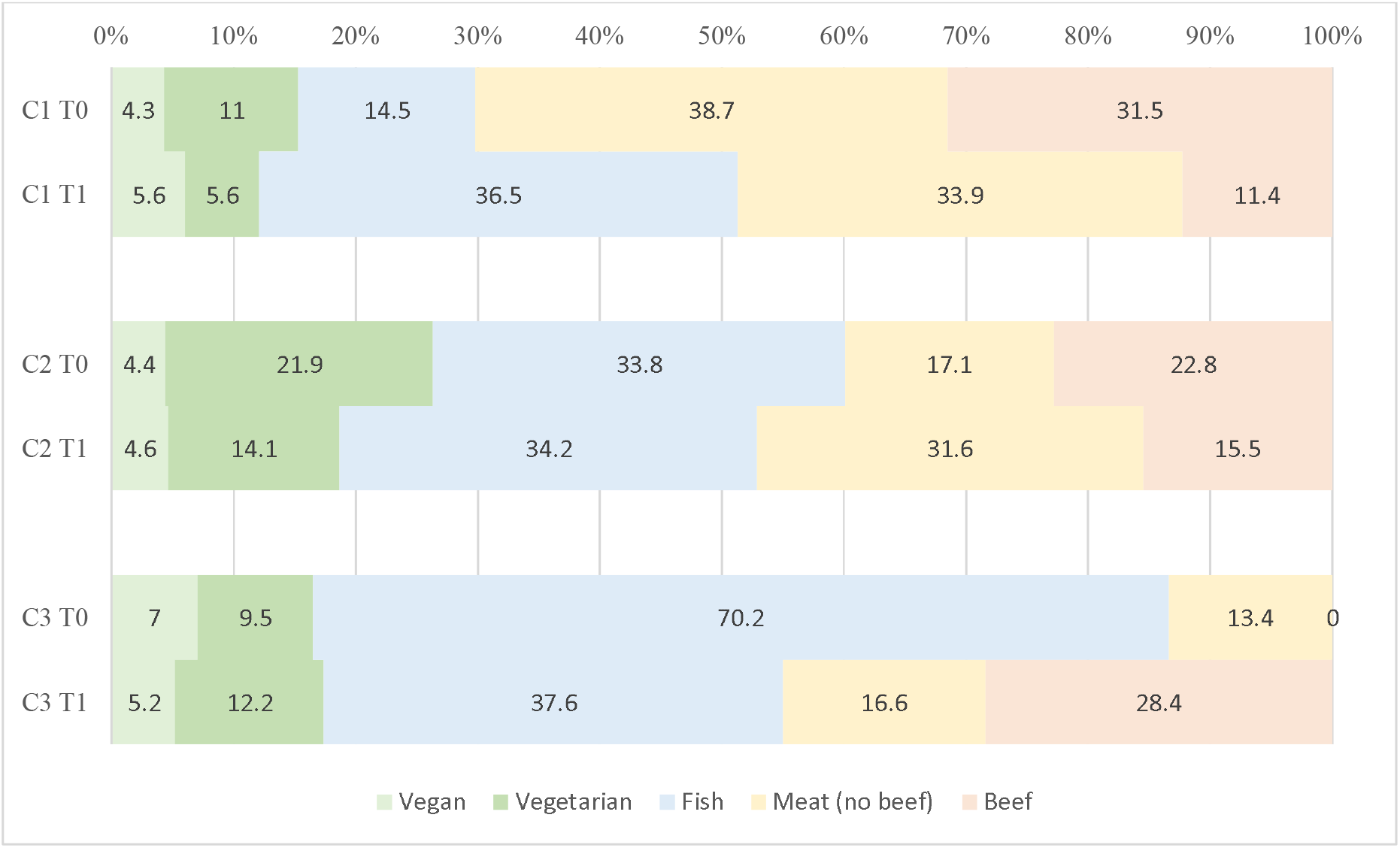
Categorization of trays in the two assessments (T0 vs. T1)

The analysis also revealed a significant association between the type of second course and the period of assessment (T0-T1) (p < 0.001). In C1, meat-based second courses (excluding beef) increased significantly (21.7% at T0 vs. 31.1% at T1), while those with beef decreased (22.4% at T0 vs. 10.2% at T1). In C2, fish second courses increased (8.8% at T0 vs. 22.1% at T1), while meat (excluding and including beef) decreased, but not significantly. In C3, meat second courses excluding beef decreased (80.4% at T0 vs. 37.6% at T1) and beef main courses increased (0% at T0 vs. 27.3% at T1). The complete results are presented in Supplementary Table 4.

As shown in Table 3, there was no significant association between fruit consumption and assessment period (T0-T1) in any of the three canteens. However, there is a significant association between side dishes and assessment time, in C1 and C3 (*Table 3*). Furthermore, in C1, where salad was switched to buffet distribution, 39.9% of users took salad at T1, compared to 23.3% at T0 (p < 0.001).

## 4 Discussion

This study provided an opportunity to evaluate the effectiveness of educational and environmental interventions in guiding users’ choices towards healthier and more sustainable habits in three northern Italian hospital canteens. The intervention varied slightly according to the spaces and logistical possibilities of each canteen. Specifically, in C1, educational posters, fruit, pulses and salad self-service, and posters with descriptive norm messages were implemented; in C2, educational posters, a reversal of the flow of dish choices starting from the side dish, and posters with descriptive norm messages were implemented; and in C3, educational posters (excluding the suggested tray composition due to catering management limitations) and posters with descriptive norm messages were implemented.

C1, the largest canteen in terms of daily user rate and managed by an external catering service, had the macronutrient profile closest to the Italian reference values (SINU Società Italiana di Nutrizione Umana. 2024). Nevertheless, users’ choices showed the worst environmental profile, probably due to the high selection of beef-containing foods and larger meals (i.e., highest median meal energy). In contrast, C3, which has the smallest user rate (and therefore the smallest sample size) and is the only canteen managed internally, showed considerable variability in results between T0 and T1. However, C3 represents, however, the canteen with the environmental profile and fibre intake closest to the reference ranges, but only at T0. Given the type of management and the resulting high variability in food offerings, only an extended observation period could provide truly representative data for this canteen. Finally, C2 is the canteen with the macronutrient profile that differs most from the Italian reference values at both assessment points. However, at T1, it has a median value for environmental indicators within the reference ranges, which may reflect the lower energy content of the meals.

### 4.1 Nutritional assessment

According to the guidelines (CREA 2018), the energy content of a lunch meal suitable for a 2000 kcal diet could be set between 700 and 800 kcal. At T0, the energy intake from the meal was below 700 kcal in C3 and above 800 kcal in C2, while at T1, the values were within the 700-800 kcal range in both canteens. However, in C1, the median energy value remained virtually unchanged and slightly above the identified range, indicating more caloric meals despite the intervention. As observed in the first assessment in 2022 (Menis et al. 2024), at T1, meals in all three canteens tended to be high in lipids. In C1 and C3, the contribution of lipids to energy intake was high but within the reference range (20-35%E) at both time points (SINU Società Italiana di Nutrizione Umana. 2024), while in C2 it was higher, with a median of 40%E at T0 and 39%E at T1. Consequently, in C2, the contribution of carbohydrates to energy was below the lower limit of the reference range (45-60%E) (SINU Società Italiana di Nutrizione Umana. 2024), with a median of 39%E at both T0 and T1. This observation confirms previous findings in studies evaluating workplace canteen settings, where meals often have an unbalanced nutritional profile, with high energy and fat content, which in turn is reflected in less healthy food choices by users (Kjøllesdal et al. 2011; Jaworowska et al. 2018). Our results confirm this imbalance in macronutrient distribution, even after the combined educational and environmental intervention focused on healthy and sustainable food choices.

At T1, the fibre content of the meal decreased compared to T0 in C2 and increased in C1 and C3; in these last two the value falls within the reference range for adults (12.6-16.7 g/1000 kcal) (SINU Società Italiana di Nutrizione Umana. 2024). One factor that may have increased fibre intake in C1 after the intervention is the partial change in service modality. At T0, users did not have autonomy in selecting fruit and vegetables, whereas at T1, pulses, salads, and fruits were offered as a buffet (notably, users often took portions of fruit, vegetables, and pulses that exceeded those allowed by canteen regulations) (Menis et al. 2024). Supporting this, after the intervention, there was an increase in trays containing salad and side dishes compared to T0. Specifically, the presence of salad rose from 23.3% to 39.9%, while that of side dishes increased from 66.5% to 71.6%. Considering this increase, it can be assumed that the combined effect of the posters raising awareness of healthy food choices and the presence of the buffet nudging choices positively impacted the selection of vegetables (Vandenbroele et al. 2020). However, in C2, where the percentage of trays containing a side dish remained unchanged, the decrease in fibre intake could be attributed to smaller portions of vegetables compared to T0 and/or the frequent presence of potatoes served and chosen as side dishes. In fact, while Italian culinary tradition often includes potatoes as a side dish, these tubers cannot be considered vegetables, as their nutrient composition is more similar to that of cereal products, which are generally consumed as part of first courses or as bread and substitutes. Moreover, the combined intervention of posters with the reversal of the flow of dish choice - starting from side dishes, salad, and fruit instead of first courses - introduced only in C2, appeared not to have had the desired effect, contrary to what would be expected from the literature on the reversal of serving flow (Graça et al. 2023). The results suggest that posters can encourage the choice of vegetables in some cases, but their effectiveness is influenced by the context in which they are implemented. The increase in vegetable choice observed in the canteen (C1), where educational posters and descriptive norm messages were combined with a buffet service offering vegetables, pulses, and fruit, supports the hypothesis that information strategies are more effective when accompanied by structural changes in food service that reduce barriers to healthy choices (e.g., allowing independent selection of combinations and quantities of vegetables) (World Health Organization 2022), compared with the same educational and descriptive norm message interventions combined with the reversal of the dish service flow. However, the increase in vegetable consumption (the percentage of trays containing a side dish rose from 63.7% to 74.5%) observed in the canteen where only educational intervention and social norms were used (C3) suggests that communication strategies can also be effective in smaller contexts independently of environmental structural changes, where vegetable options are probably already perceived as appealing. Indeed, a positive effect of educational posters and descriptive norm messages without other environmental changes was previously highlighted by other authors (Thomas et al. 2017; Collins et al. 2019). Social norms may have been particularly influential in this setting, as the small size of the canteen facilitate familiarity and cohesion among users, amplifying peer influence on food choices (Rathbone et al. 2023). The combined scenario (appealing healthy options and cohesive social groups) may have amplified the impact of the educational and descriptive norm message intervention, confirming that the effectiveness of nutritional communication tools depends not only on the content itself, but also on how they are integrated with the physical and social environment of the meal (Çoker et al. 2022). On the contrary, as previously noted, the absence of significant changes in the canteen where educational materials and norm messages were associated with reversing the order of dishes suggests that the effectiveness of nudges is highly dependent on the setting and the offer. Unlike what is reported in this study, evidence highlights the effectiveness of reversing the order of dishes (Graça et al. 2023), presenting the healthiest ones first or those that are intended to be encouraged. Recent studies, such as those by Hunninghaus (Hünninghaus et al. 2025) and Lohmann (Lohmann et al. 2024), show that rearranging the order of dishes, particularly by placing the healthiest options first on the menu, significantly increases their selection. In this study, the opposite outcome could depend on the context: users may have had more established eating habits and therefore been less sensitive to environmental changes; furthermore, limited meal time, and eating in groups may have reduced their attention and ability to adopt healthier food choices. As reported in a recent systematic review, these factors can hinder change and affect people’s decision-making abilities (Deslippe et al. 2023). Individual and contextual factors such as established food preferences and socioeconomic and cultural barriers hinder the transition to more sustainable food choices (Thornton et al. 2013; Viroli et al. 2023) that improve health and reduce the overall ecological footprint (Viroli et al. 2023; Kalmpourtzidou et al. 2025). In conclusion, regarding the effectiveness of educational and descriptive norm message interventions and the buffet serving modality, the results suggest a role for these interventions in encouraging an increase in the selection of salad and side dishes, while they did not produce significant changes in the choice of fruit. This difference could be explained by the fact that fruit and vegetables, although often considered together in nutritional recommendations, respond to different behavioral determinants. Vegetables are generally perceived as a structural component of the main meal (Bel-Serrat et al. 2022), and their choice can be more easily influenced by messages promoting healthy meals. In contrast, fruit is frequently associated with optional consumption, such as dessert or snacks (Carroll et al. 2024), and is more influenced by individual preferences and established habits (Wellard-Cole et al. 2023). Furthermore, fruit may require greater behavioral effort (i.e. peelling, slicing, eating by hand) than ready-to-eat vegetables, reducing the effectiveness of purely educational interventions (Wellard-Cole et al. 2023). These results suggest that improving fruit consumption in hospital canteens may require more than just communication strategies, including, for example, more targeted environmental interventions such as increasing visibility, variety, and ease of access to the product (e.g., pre-cut fruit), as highlighted in the systematic review by Metcalfe J et al. (Metcalfe et al. 2020).

### 4.2 Sustainability assessment

It is noteworhty that in C1 and C2, the median carbon footprint decreased after the intervention (from 1338 g CO_2_ eq./meal to 1227 g CO_2_ eq./meal in C1 and from 1094 g CO_2_ eq./meal to 966 g CO_2_ eq./meal in C2). However, in C3, the median carbon footprint increased (from 773 g CO_2_ eq./meal to 1126 g CO_2_ eq./meal). In a previous study on pre-intervention assessment, researchers highlighted that dishes containing beef have the highest value of carbon footprint (Menis et al. 2024), as beef is the food with the greatest environmental impact (BCFN 2018). It is estimated that 100g of beef corresponds to approximately 2.5 kg CO_2_ eq, which is more than double the upper limit of the reference range for one complete meal (BCFN 2018). In C1 and C2, after the intervention, the number of trays and second courses containing beef decreased, likely contributing to the reduction in carbon footprint from T0 to T1 in both canteens. This reduction is further supported by the decrease in the selection of cheese as a second course, which, being derived from beef, contributes significantly to the carbon footprint. The choice of cheese decreased from 12.4% to 7.2% in C1 and from 14.5% to 12.9% in C2. This suggests that environmental interventions, combined with descriptive norm messages and educational strategies such as food pyramids, healthy plate composition, and recommended daily menus, may have increased users’ awareness and led to more informed choices. In C3, however, the carbon footprint increased, despite a significant decrease in the number of second courses containing meat (from 80.4% to 37.6%). This result can be explained by the absence of beef at T0, whereas at T1, during the week of analysis, two beef options were introduced among the second courses and one among the first courses (Thornton et al. 2013; Viroli et al. 2023; Kalmpourtzidou et al. 2025). Overall, all canteens have high carbon footprint and water footprint values; therefore, alongside nudging interventions, it is necessary to improve the food offered to make it more appealing to users, who report that making healthy choices is difficult due to the limited availability and quality of healthy options (Basilico et al. 2025).

### 4.3 Strengths and limitations

The main strength of this study is the analysis of more than 2,850 actual meal choices across multiple hospital canteens, with a comparison of choices before and after combined interventions focused on educational and environmental prompts to identify valuable and suitable interventions in specific contexts. The results contribute to expanding the available data on the most effective types of intervention in collective catering for adults. As reported in the previous pre-intervention study (Menis et al. 2024), the evaluation of trays using photographic documentation enabled the identification of any non-standardized portions, allowing for a more accurate estimate of nutritional and sustainability variables. Additionally, no recipes from the literature were used; information on portions and preparation was provided directly by the staff of the individual canteens. However, for a correct interpretation of the comparison between T0 and T1 for each canteen, it is necessary to consider that the differences found between T0 and T1 may be partially due to variations in daily menus. In C2, there was a rotational menu that changed weekly over eight weeks; in C3, the menu was extremely variable, as it was an internally managed canteen without a pre-decided menu; while in C1, there were only two menu options per season that alternated. Therefore, although the season of analysis was the same in both assessments (August and September), the dishes offered were not the same. Moreover, regarding nutritional data, it is necessary to consider that the analysis is limited to the lunch meal and therefore does not allow determination of whether any nutritional imbalance found in this meal is also reflected in the users’ overall diet. Other limitations are reported in the pre-intervention study (Menis et al. 2024), such as: choices varied depending on the time (some dishes ran out earlier), the evaluation of trays was only visual and therefore estimated, and the analysis of CF and WF was based on an environmental database derived from the literature, which requires continuous updates. Finally, it was not possible to implement a uniform intervention for all canteens, as it was tailored to each individual setting; furthermore, it was not possible to estimate its effectiveness, as the pre- and post-intervention samples were similar but not statistically comparable.

## 5 Conclusions

Even after the intervention, the dishes chosen by users tended to be high in lipids and low in carbohydrates in terms of their contribution to energy intake, compared to the Italian dietary reference values. The different dishes offered in the two observation periods did not allow a robust comparison before and after the intervention in each canteen. In the larger, externally managed canteen (C1), the presence of salad, fruit and pulses in buffet style, along with descriptive norm messages and educational posters may have contributed to a higher fibre intake after the intervention. However, in the small externally managed canteen (C2) the unbalanced macronutrient profile, and the decrease in fibre content from T0 to T1, did not align with the expected increase in vegetable or fruit intake, suggesting a lack of effectiveness of the communicative elements combined with the reverse flow of dish choice starting from vegetables and fruit. From a sustainability perspective, the presence or absence of dishes containing beef within the food offer (observed in C3) was confirmed as a crucial variable in determining the carbon footprint of users’ choices. Future strategies to improve nutritional composition and sustainability in hospital canteens should therefore combine actions to raise awareness with structural changes in the food offer. Indeed, canteens play a key role in providing options that meet nutritional and sustainability criteria. Moreover, future studies should also investigate the perception of the visual appeal and palatability of healthier food choices, as healthier and more sustainable options should not only be available but also desirable for users to select.

Although only modest results were obtained through the intervention based on food choices, hospital canteens remain favorable settings for promoting educational initiatives, thanks to the high volume of meals served and the social and cultural role of hospitals and healthcare workers in promoting healthy and sustainable lifestyles. In this context, encouraging a balanced diet not only supports the prevention of chronic diseases and can help reduce indirect healthcare costs in the long term, but also reflects a broader organizational vision that considers employee health a priority, promoting a healthy and cohesive working community.

## 6 Declarations

### 6.1 Author Contributions

LB, EM, DM, FF, LG, FC, AC, MM, LL, ES, AC and MP designed the research; DM, FF, PC, DZ and CLG collected data; LB, EM, DM, FF and MP discussed investigation methodology and contributed to result interpretation; EM, DM, FF and MD performed data analysis; LB and MP supervised the study conduction; EM, FF and DM wrote the original draft; LB and MP revised contents; all authors revised the paper and agreed with the final version of the manuscript.

### 6.2 Funding

This research did not receive any specific grant from funding agencies in the public, commercial, or not-for-profit sectors.

### 6.3 Ethics approval and consent to participate

The study was conducted in accordance with the Declaration of Helsinki, and approved by the Institutional Review Board of the University of Udine (proto-col code 108/2022, date of approval: July 6th 2022, amendment protocol code 122/2023, date of ap-proval: July 3rd 2023). Informed consent was obtained from all subjects involved in the study.

### 6.4 Availability of data and materials

All data were processed in full compliance with European data protection law (GDPR EU No. 2016/679) and Data Protection Regulation (EU) 2018/1725. Data will be made available by the authors on request.

### 6.5 Declarations of competing interest

The authors declare that they have no known competing financial interests or personal relationships that could have appeared to influence the work reported in this paper.

## Notes

### Competing Interest Statement

The authors have declared no competing interest.

### Author Declarations

The Institutional Review Board of the University of Udine (proto-col code 108/2022, date of approval: July 6th 2022, amendment protocol code 122/2023, date of ap-proval: July 3rd 2023) gave ethical approval for this work.

