## Supplementary Materials for "Sustainability and nutritional composition of food choices in hospital canteens: a pre-post intervention study"

**Supplementary Table 1. Results of the post-hoc Dunn test analysis with Bonferroni adjustment on energy, nutrients and sustainability variables between the choices in C1, C2 and C3 referred to T1**

| **Variables** | **C2 vs. C3^c^** | **C1 vs. C2^c^** | **C1 vs. C3^c^** |
| --- | --- | --- | --- |
| **Energy (kcal/tray)** | 1.000 | <0.001 | <0.001 |
| **Protein (%E)** | 0.0049 | 0.030 | 1.000 |
| **Lipids (%E)** | <0.001 | <0.001 | <0.001 |
| **Carbohydrates (%E)** | 0.454 | <0.001 | <0.001 |
| **Fibre (%E)** | <0.001 | <0.001 | <0.001 |
| **Sugars (%E)** | <0.001 | <0.001 | 1.000 |
| **SFA (%E)** | 1.000 | <0.001 | <0.001 |
| **MUFA (%E)** | 0.437 | <0.001 | <0.001 |
| **PUFA (%E)** | <0.001 | <0.001 | 0.167 |
| **EPA + DHA (mg)** | 1.000 | <0.001 | <0.001 |
| **Protein (g)** | 0.048 | <0.001 | <0.001 |
| **Lipids (g)** | 0.052 | <0.001 | 0.960 |
| **Carbohydrates (g)** | 1.000 | 0.001 | <0.001 |
| **Sugars (g)** | <0.001 | <0.001 | 0.007 |
| **Fibre (g)** | <0.001 | <0.001 | 0.874 |
| **SFA (g)** | 1.000 | 1.000 | 1.000 |
| **MUFA (g)** | 0.154 | 1.000 | 0.009 |
| **PUFA (g)** | <0.001 | <0.001 | 0.067 |
| **CF (g CO_2_eq./tray)** | <0.001 | <0.001 | 0.026 |
| **WF (L H_2_O/tray)** | <0.001 | <0.001 | 1.000 |

Notes: ^c^ Post-hoc Dunn test with Bonferroni adjustement

**Supplementary Table 2. Comparison between T0 and T1 for nutritional values and sustainability food choices in the three contexts (C1, C2, C3). The median increased in T1 is shown in green, while the median decreased in T1 is shown in red.**

| **Variables** | | **C1 at T0 vs. C1 at T1** | | **C2 at T0 vs. C2 at T1** | **C3 at T0 vs. C3 at T1** |
| --- | --- | --- | --- | --- | --- |
| **Energy (kcal/tray)** |  | | < .001 | < .001 | < .001 |
| **Protein (%E)** |  | | < .001 | 0.001 | 0.119 |
| **Lipids (%E)** |  | | 0.002 | 0.806 | 0.004 |
| SFA (%E) |  | | < .001 | 0.009 | 0.015 |
| MUFA (%E) |  | | < .001 | 0.067 | < .001 |
| PUFA (%E) |  | | < .001 | < .001 | 0.016 |
| **Carbohydrates (%E)** |  | | 0.639 | 0.265 | < .001 |
| Sugars (%E) |  | | 0.037 | 0.003 | 0.708 |
| **Fibre (%E)** |  | | < .001 | 0.069 | 0.002 |
| **Protein (g)** |  | | 0.083 | 0.030 | < .001 |
| **Lipids (g)** |  | | 0.743 | < .001 | < .001 |
| SFA (g) |  | | < .001 | < .001 | 0.282 |
| MUFA (g) |  | | < .001 | < .001 | < .001 |
| PUFA (g) |  | | 0.078 | 0.869 | < .001 |
| EPA + DHA (mg) |  | | < .001 | 0.021 | < .001 |
| **Carbohydrates (g)** |  | | 0.002 | < .001 | 0.172 |
| Sugars (g) |  | | 0.759 | < .001 | 0.061 |
| **Fibre (g)** |  | | < .001 | < .001 | < .001 |
| **CF (g CO2eq./tray)** |  | | < .001 | < .001 | < .001 |
| **WF (L H2O/tray)** |  | | 0.107 | < .001 | < .001 |
| Notes: C1, canteen 1; C2, canteen 2; C3, canteen 3; SFA, saturated fatty acids; MUFA, monounsaturated fatty acids; PUFA, polyunsaturated fatty acids; EPA, eicosapentaenoic acid; DHA, docosahexaenoic acid; CF, carbon footprint; WF, water footprint;  *Mann-Whitney U test |  | | |  |  |

**Supplementary Table 3. Absolute and relative frequency of tray categories in T0 and T1 in the three canteens (C1, C2, C3) and respective Pearson standardized residuals**

| **Type of tray in C1** |  | **T0** | **T1** | | **Total** | **p-value** |
| --- | --- | --- | --- | --- | --- | --- |
|  |  |  |  |  | | <0.001^b^ |
| **Vegan** | Count | 34 (4.3) | 56 (5.6) | 90 | |  |
|  | Standardized residuals | -1.270 | 1.270 |  | |  |
| **Vegetarian** | Count | 88 (11) | 126 (12.5) | 214 | |  |
|  | Standardized residuals | -0.985 | 0.985 |  | |  |
| **Fish** | Count | 116 (14.5) | 367 (36.5) | 483 | |  |
|  | Standardized residuals | -10.468 | 10.468 |  | |  |
| **Meat (no beef)** | Count | 309 (38.7) | 341 (33.9) | 650 | |  |
|  | Standardized residuals | 2.105 | -2.105 |  | |  |
| **Beef** | Count | 251 (31.5) | 115 (11.4) | 366 | |  |
|  | Standardized residuals | 10.492 | -10.492 |  | |  |
| **Total** | Count | 798 | 1.005 | 1.803 | |  |
| **Type of tray in C2** |  | **T0** | **T1** | **Total** | | **p-value** |
|  |  |  |  |  | | <0.001^b^ |
| **Vegan** | Count | 10 (4.4) | 16 (4.6) | 26 | |  |
|  | Standardized residuals | -0.120 | 0.120 |  | |  |
| **Vegetarian** | Count | 50 8 (21.9) | 49 (14.1) | 99 | |  |
|  | Standardized residuals | 2.442 | -2.442 |  | |  |
| **Fish** | Count | 77 (33.8) | 119 (34.2) | 196 | |  |
|  | Standardized residuals | -0.105 | 0.105 |  | |  |
| **Meat (no beef)** | Count | 39 (17.1) | 110 (31.6) | 149 | |  |
|  | Standardized residuals | -3.887 | 3.887 |  | |  |
| **Beef** | Count | 52 (28.8) | 54 (15.5) | 106 | |  |
|  | Standardized residuals | 2.208 | -2.208 |  | |  |
| **Total** | Count | 228 | 348 | 576 | |  |
| **Type of tray in C3** |  | **T0** | **T1** | **Total** | | **p-value** |
|  |  |  |  |  | | <0.001^b^ |
| **Vegan** | Count | 14 (7) | 14 (5.2) | 28 | |  |
|  | Standardized residuals | 0.818 | -0.818 |  | |  |
| **Vegetarian** | Count | 19 (9.5) | 33 (12.2) | 52 | |  |
|  | Standardized residuals | -0.935 | 0.935 |  | |  |
| **Fish** | Count | 141 (70.2) | 102 (37.6) | 243 | |  |
|  | Standardized residuals | 6.988 | -6.988 |  | |  |
| **Meat (no beef)** | Count | 27 (13.4) | 45 (16.6) | 72 | |  |
|  | Standardized residuals | -0.948 | 0.948 |  | |  |
| **Beef** | Count | 0 | 77 (28.4) | 77 | |  |
|  | Standardized residuals | -8.261 | 8.261 |  | |  |
| **Total** | Count | 201 | 271 | 472 | |  |

Notes: The values are presented as absolute frequencies and column percentages; ^b^ = Chi-squared test

**Supplementary Table 4. Absolute and relative frequency of second course categories in T0 and T1 in the three canteens (C1, C2, C3) and respective Pearson standardized residuals**

| **Second course in C1** |  | | **T0** | | | **T1** | **Total** | **p-value^b^** |
| --- | --- | --- | --- | --- | --- | --- | --- | --- |
|  |  |  | |  |  |  |  | <0.001 |
| **0** |  | Count | |  | 121(15.2) | 204 (20.3) | 325 |  |
|  |  | Standardized residuals | |  | -2.818 | 2.818 |  |  |
| **Vegan** |  | Count | |  | 0 | 23 (2.3) | 23 |  |
|  |  | Standardized residuals | |  | -4.301 | 4.301 |  |  |
| **Vegetarian** |  | Count | |  | 99 (12.4) | 72 (7.2) | 171 |  |
|  |  | Standardized residuals | |  | 3.773 | -3.773 |  |  |
| **Meat (no beef)** |  | Count | |  | 173 (21.7) | 313 (31.1) | 486 |  |
|  |  | Standardized residuals | |  | -4.499 | 4.499 |  |  |
| **Fish** |  | Count | |  | 226 (28.3) | 291 (29) | 517 |  |
|  |  | Standardized residuals | |  | -0.296 | 0.296 |  |  |
| **Beef** |  | Count | |  | 179 (22.4) | 102 (10.2) | 281 |  |
|  |  | Standardized residuals | |  | 7.141 | -7.141 |  |  |
| **Total** |  | Count | |  | 798 | 1.005 | 1.803 |  |
| **Second course in C2** |  |  | |  | **T0** | **T1** | **Total** | **p-value^b^** |
|  |  |  | |  |  |  |  | <0.001 |
| **0** |  | Count | |  | 39 (17.1) | 36 (10.3) | 75 |  |
|  |  | Standardized residuals | |  | 2.358 | -2.358 |  |  |
| **Vegan** |  | Count | |  | 9 (4) | 26 (7.5) | 35 |  |
|  |  | Standardized residuals | |  | -1.731 | 1.731 |  |  |
| **Vegetarian** |  | Count | |  | 33 (14.5) | 45 (12.9) | 78 |  |
|  |  | Standardized residuals | |  | 0.529 | -0.529 |  |  |
| **Meat (no beef)** |  | Count | |  | 96 (42.1) | 131 (37.6) | 227 |  |
|  |  | Standardized residuals | |  | 1.072 | -1.072 |  |  |
| **Fish** |  | Count | |  | 20 (8.8) | 77 (22.1) | 97 |  |
|  |  | Standardized residuals | |  | -4.188 | 4.188 |  |  |
| **Beef** |  | Count | |  | 31 (13.6) | 33 (9.5) | 64 |  |
|  |  | Standardized residuals | |  | 1.536 | -1.536 |  |  |
| **Total** |  | Count | |  | 228 | 348 | 576 |  |
| **Second course in C3** |  |  | |  | **T0** | **T1** | **Total** | **p-value^b^** |
|  |  |  | |  |  |  |  | <0.001 |
| **0** |  | Count | |  | 0 | 42 (15.5) | 42 |  |
|  |  | Standardized residuals | |  | -5.049 | 5.049 |  |  |
| **Vegan** |  | Count | |  | 1 (0.7) | 9 (3.3) | 10 |  |
|  |  | Standardized residuals | |  | -1.696 | 1.696 |  |  |
| **Vegetarian** |  | Count | |  | 13 (8.8) | 11 (4.1) | 24 |  |
|  |  | Standardized residuals | |  | 1.989 | -1.989 |  |  |
| **Meat (no beef)** |  | Count | |  | 119 (80.4) | 102 (37.6) | 221 |  |
|  |  | Standardized residuals | |  | 8.381 | -8.381 |  |  |
| **Fish** |  | Count | |  | 15 (10.1) | 33 (12.2) | 48 |  |
|  |  | Standardized residuals | |  | -0.627 | 0.627 |  |  |
| **Beef** |  | Count | |  | 0 | 74 (27.3) | 74 |  |
|  |  | Standardized residuals | |  | -7.006 | 7.006 |  |  |
| **Total** |  | Count | |  | 148 | 271 | 419 |  |

Notes: The values are presented as absolute frequencies and column percentages; ^b^ = Chi-squared test

**Supplementary Figure 1. Distribution of nutritional and sustainability values in C1 at T0 and T1**

a) **Energy (kcal/tray)**

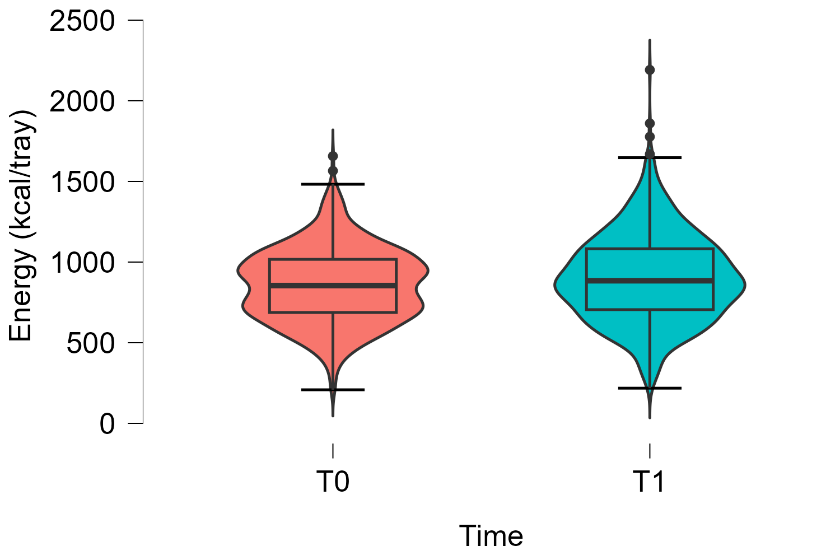

b) **Protein (%E)**

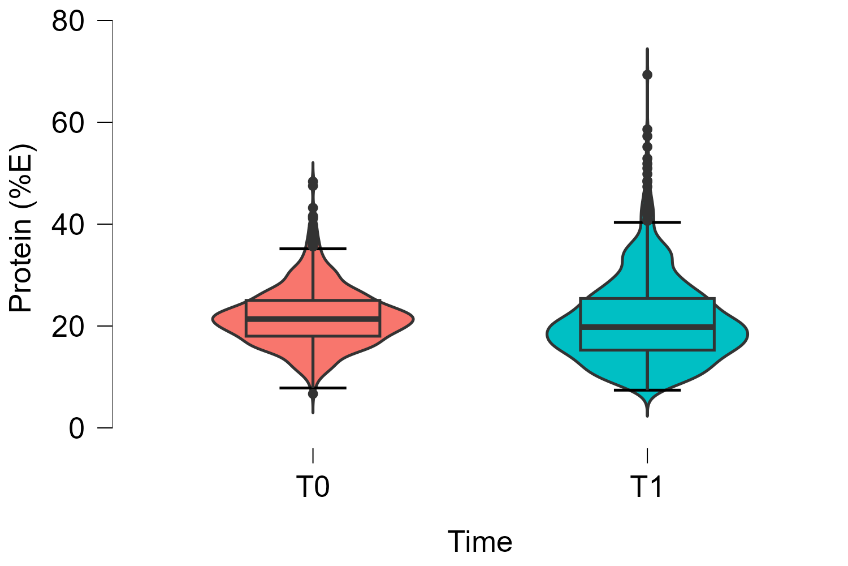

c) **Lipids (%E)**

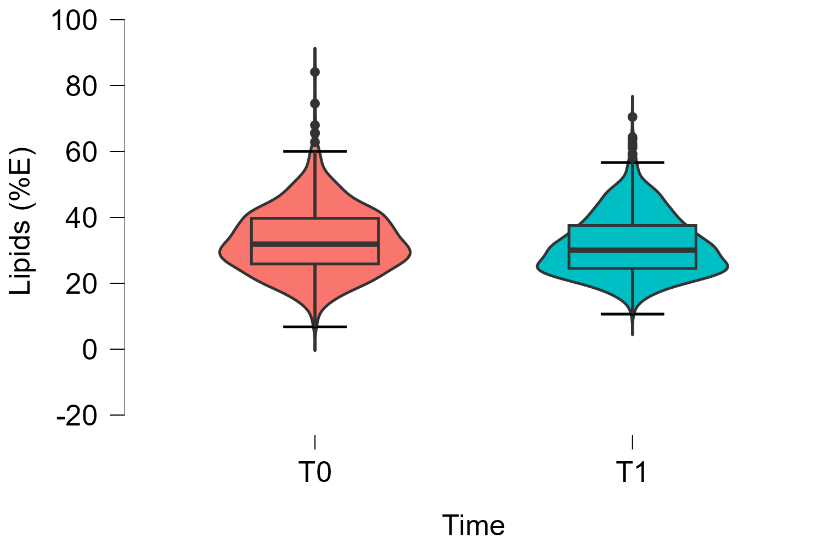

d) **Carbohydrates (%E)**

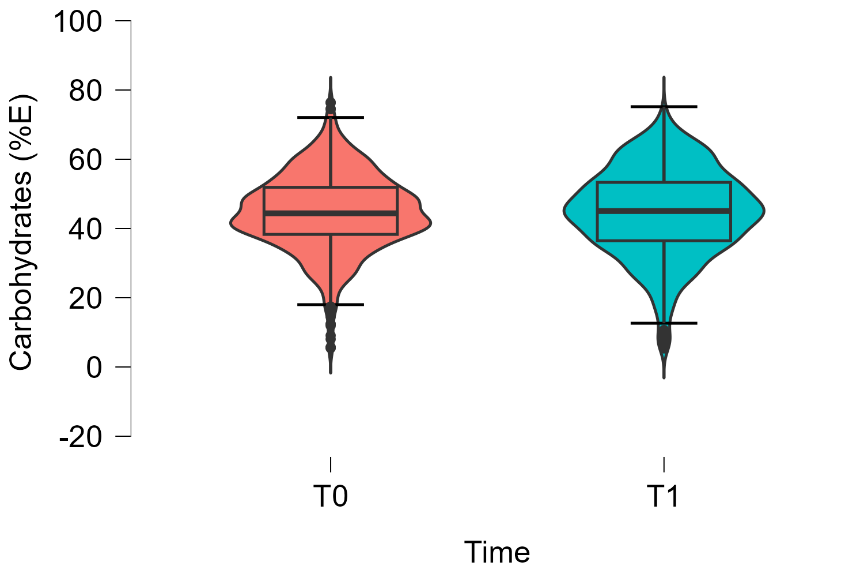

e) **Fibre (%E)**

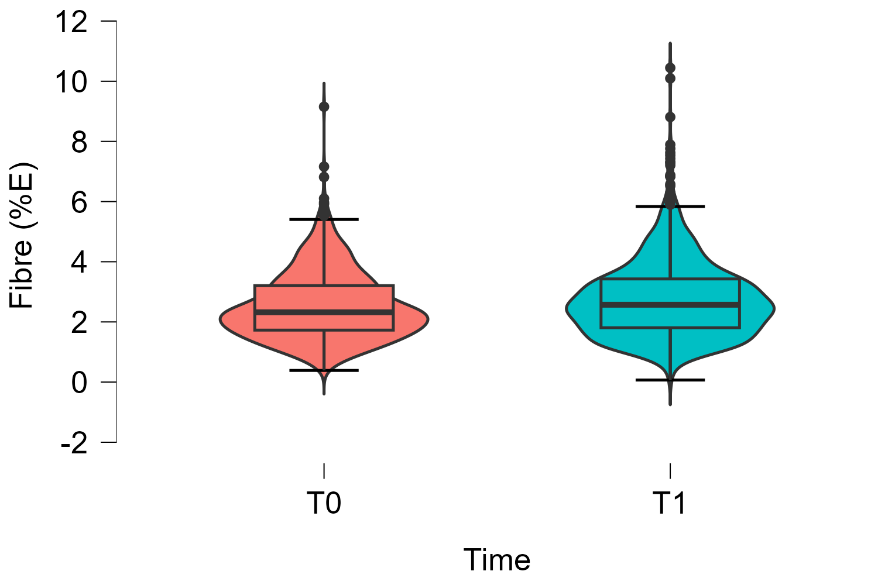

f) **Sugars (%E)**

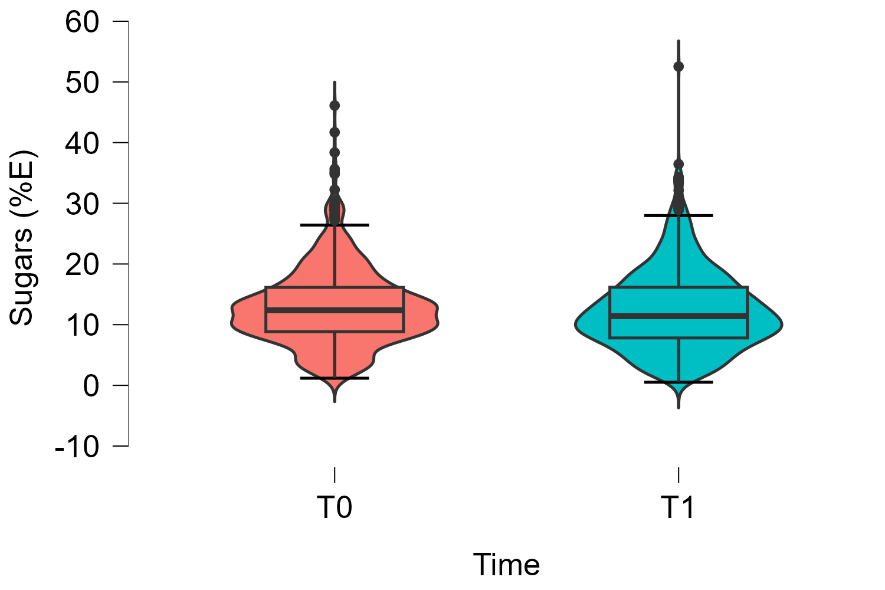

g) **SFA (%E)**

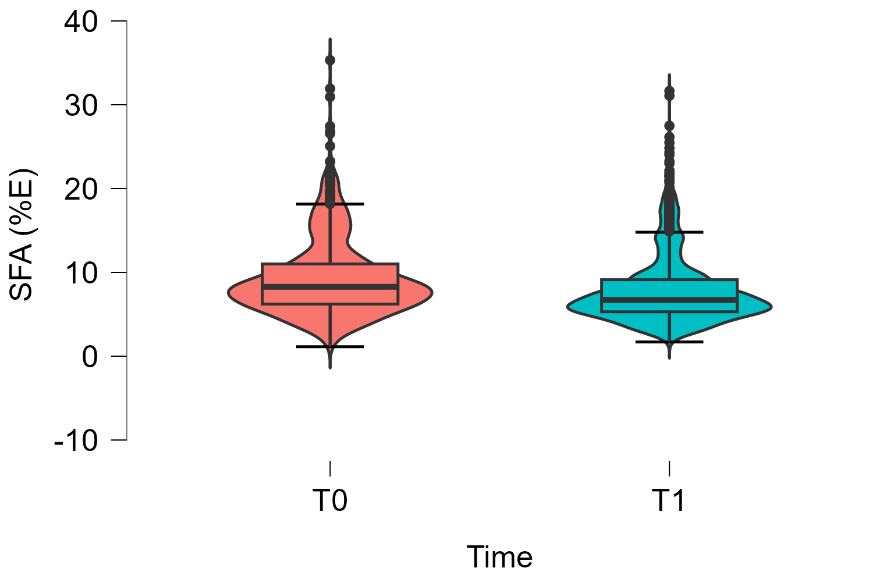

h) **MUFA (%E)**

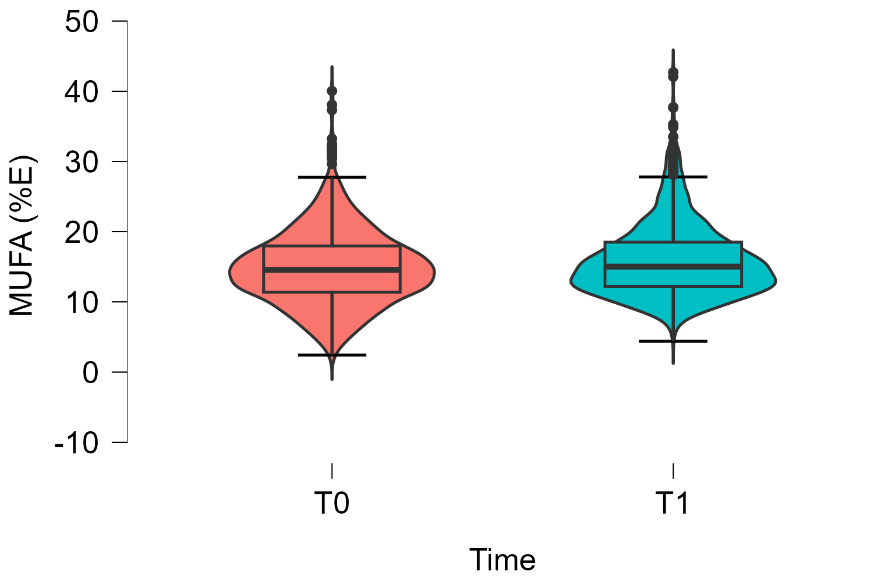

i) **PUFA (%E)**

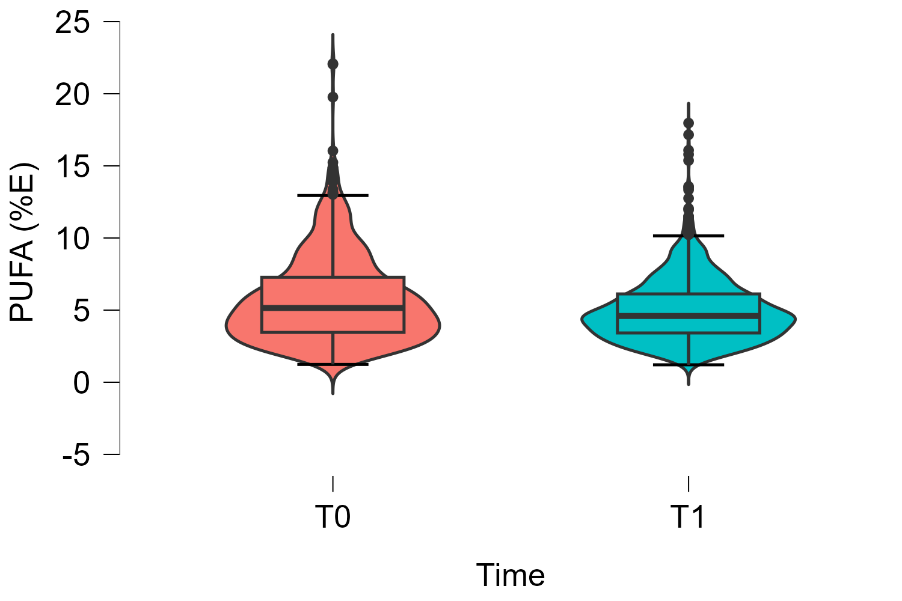

j) **EPA + DHA (mg)**

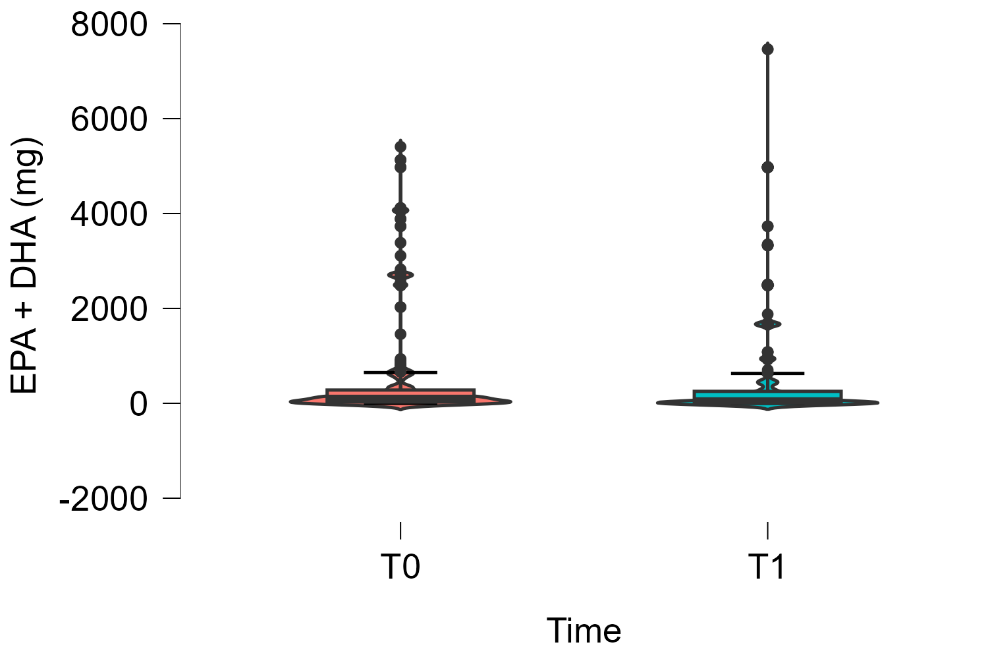

k) **Protein (g)**

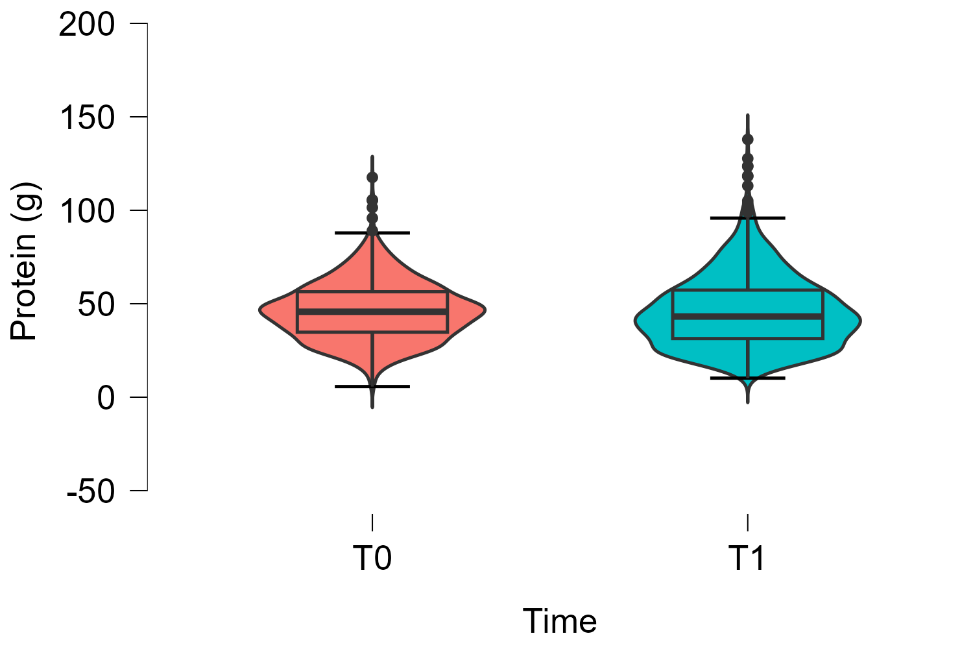

l) **Lipids (g)**

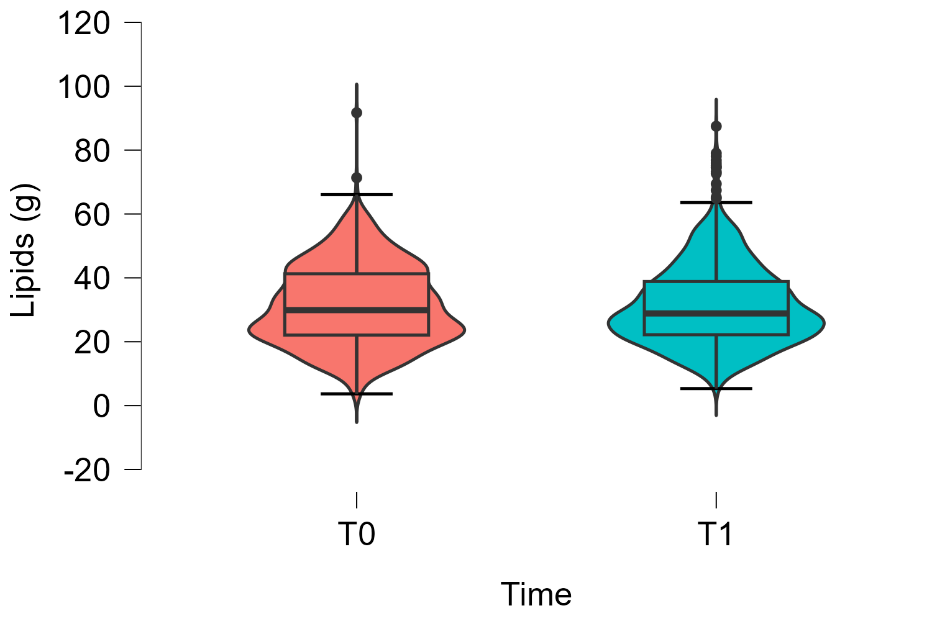

m) **Carbohydrates (g)**

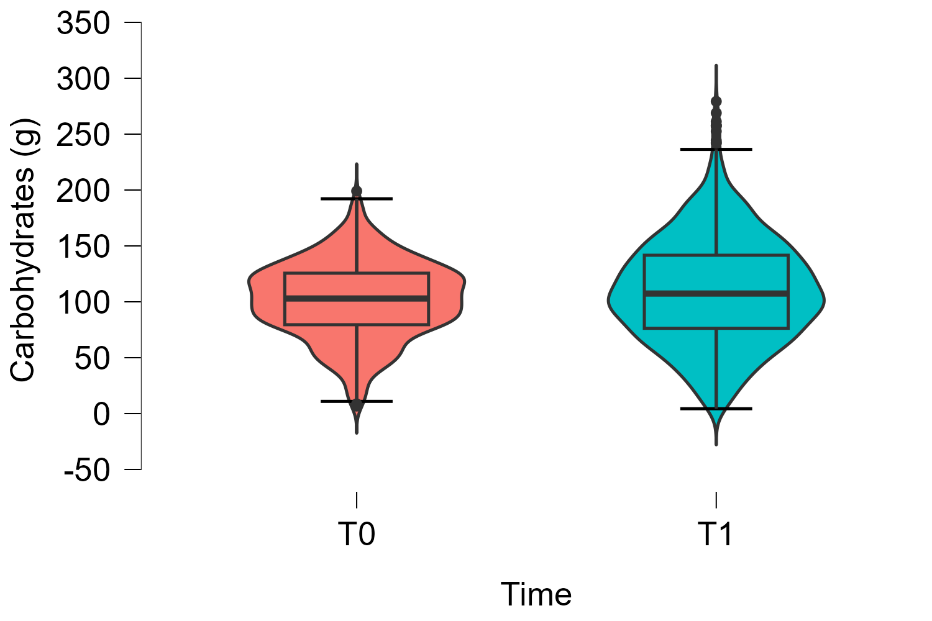

n) **Sugars (g)**

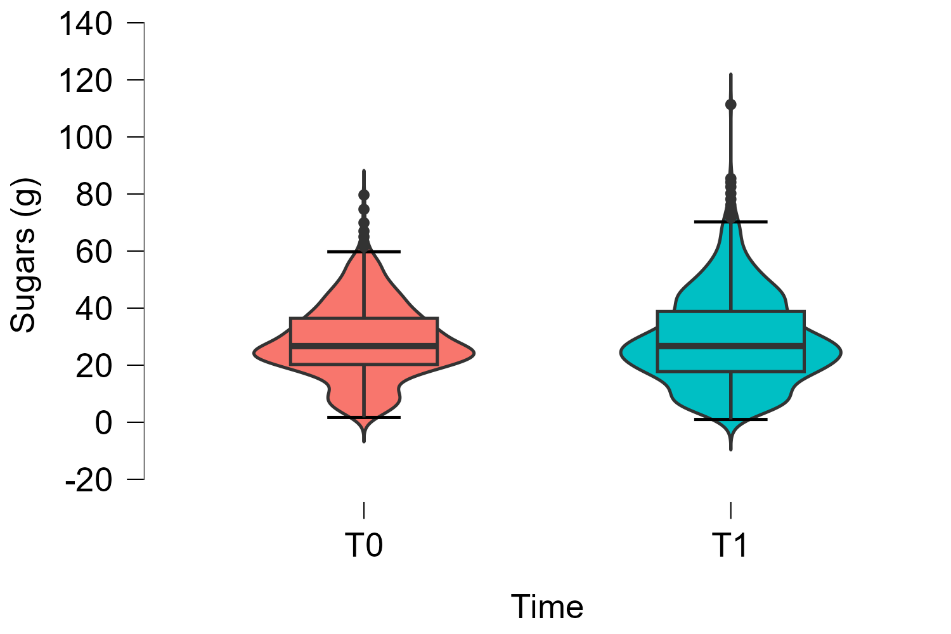

o) **Fibre (g)**

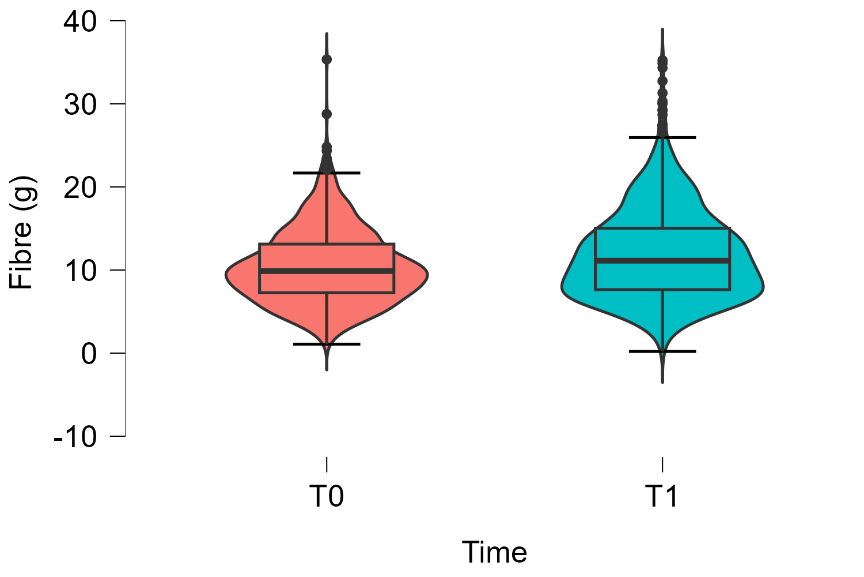

p) **SFA (g)**

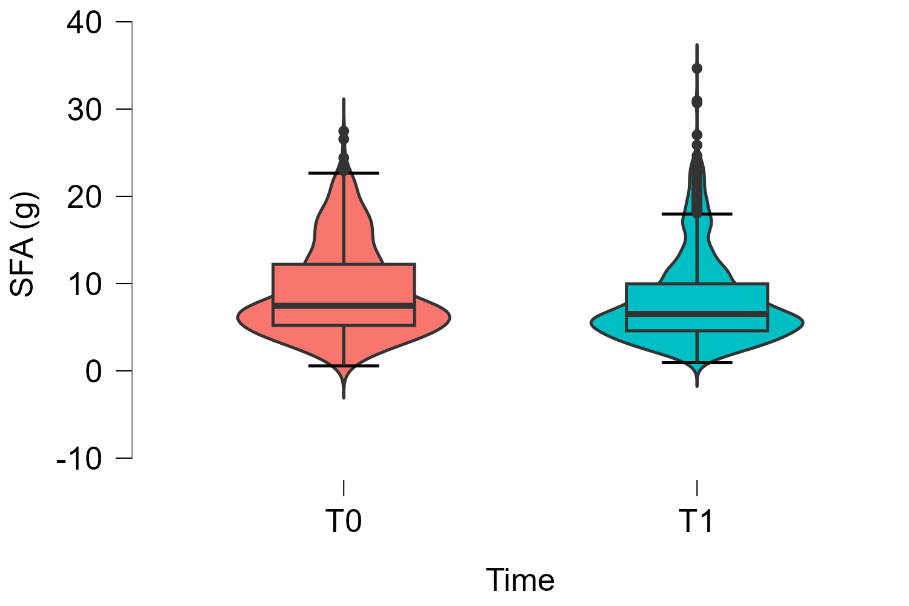

q) **MUFA (g)**

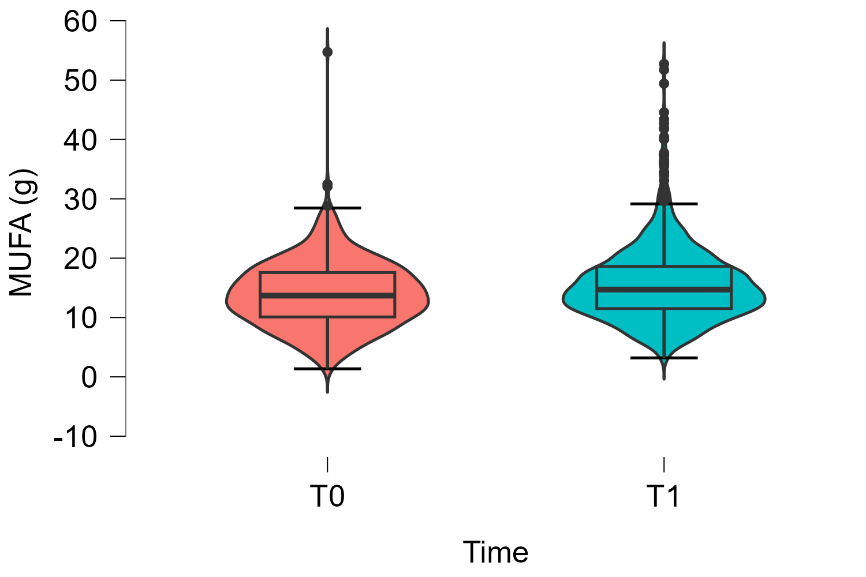

r) **PUFA (g)**

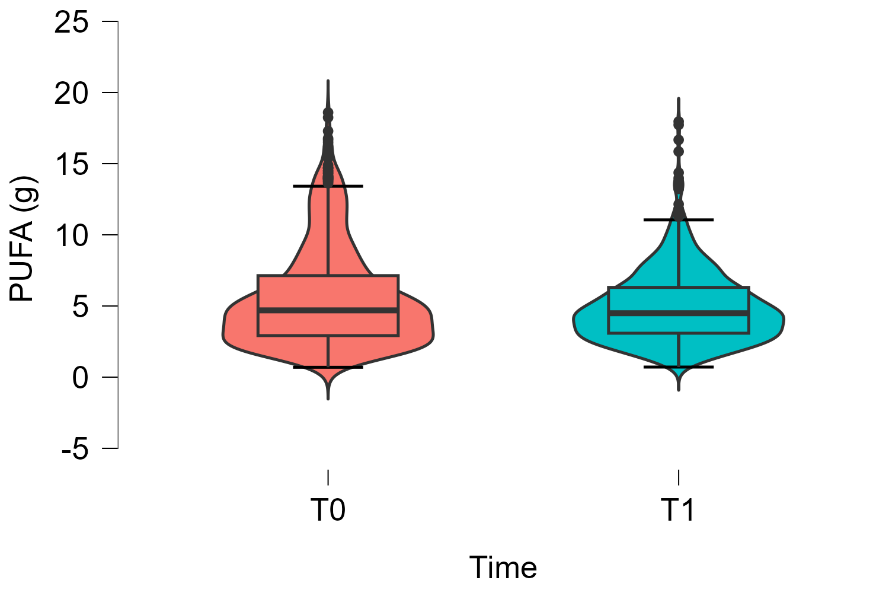

s) **CF (g CO2eq./tray)**

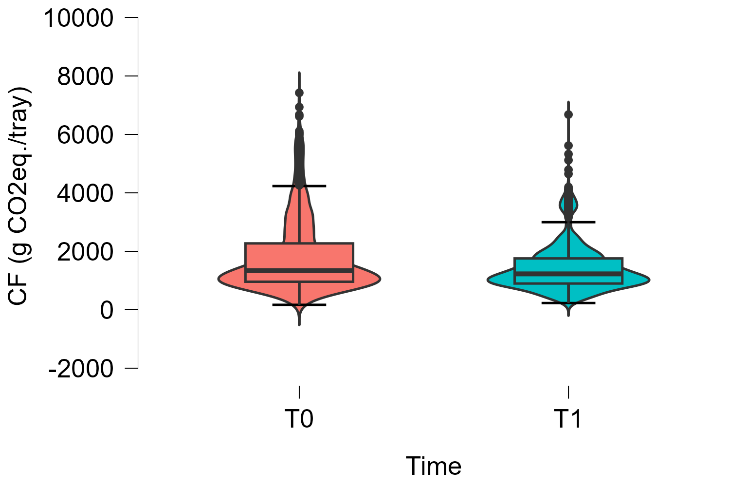

#### t) **WF (L H2O/tray)**

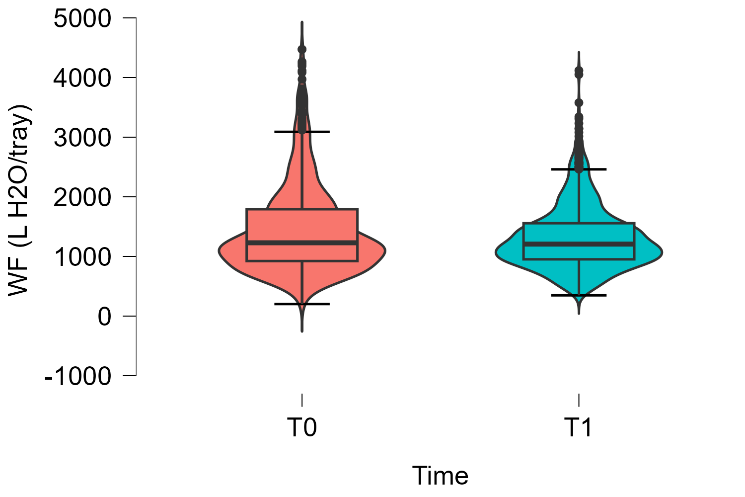

**Supplementary Figure 2. Distribution of nutritional and sustainability values in C2 at T0 and T1**

a) **Energy (kcal/tray)**

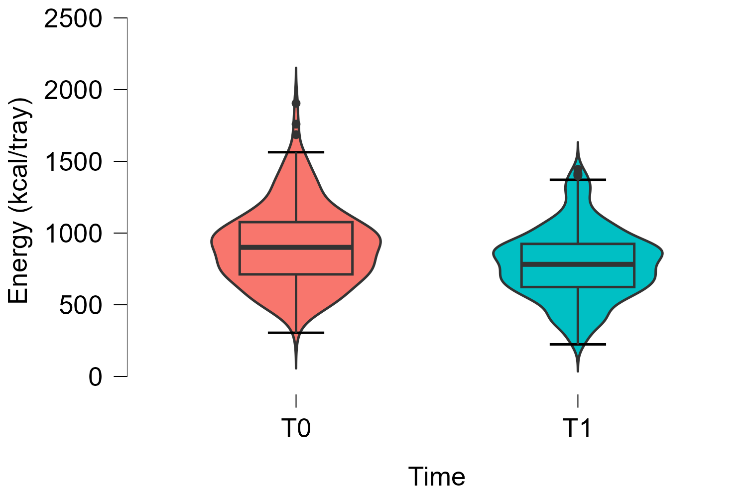

b) **Protein (%E)**

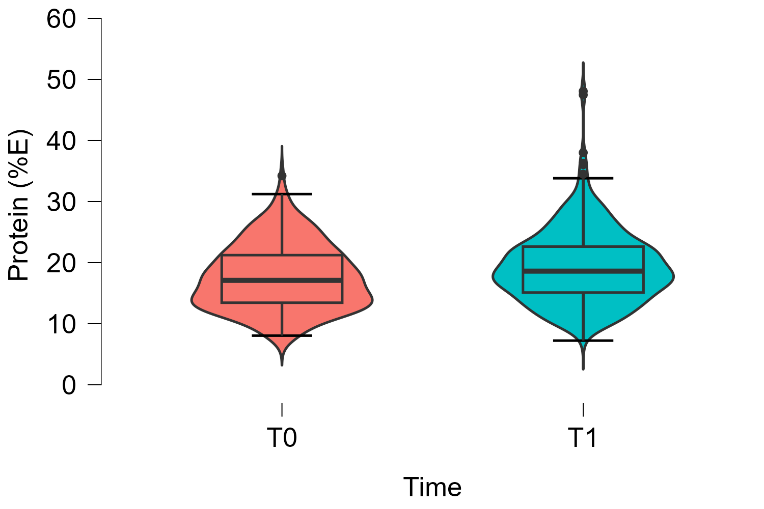

c) **Lipids (%E)**

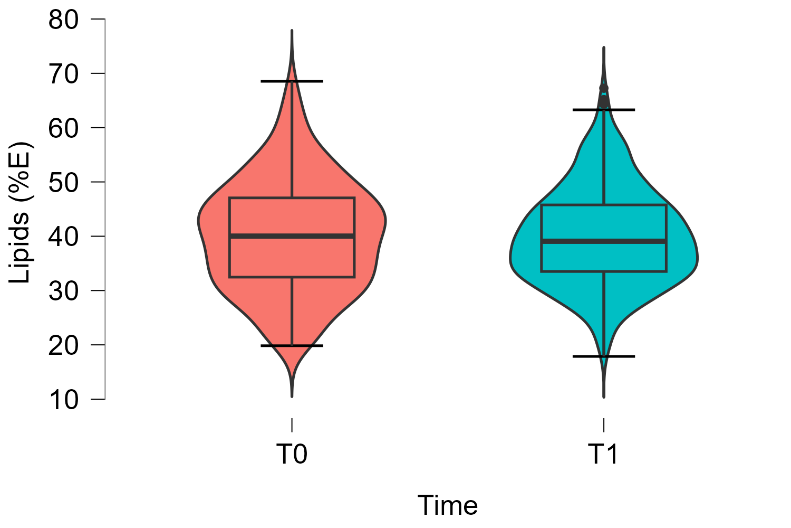

d) **Carbohydrates (%E)**

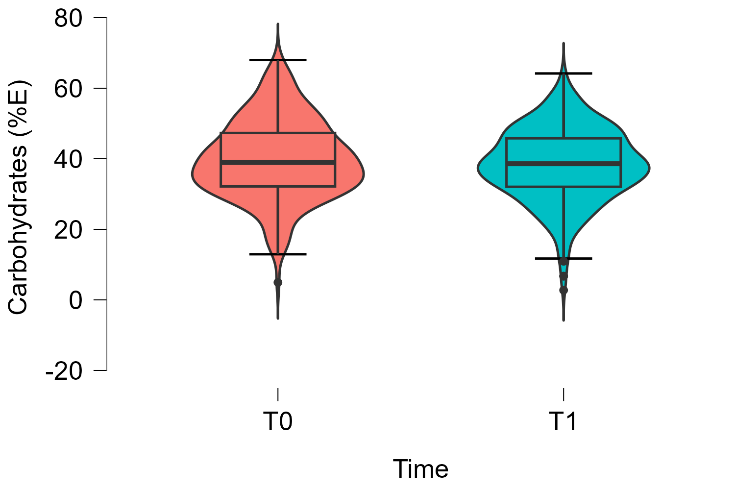

e) **Fibre (%E)**

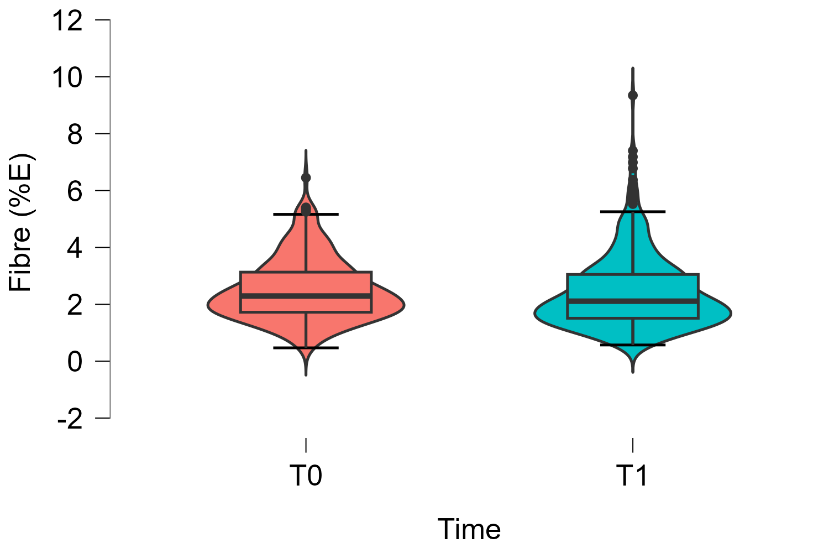

f) **Sugars (%E)**

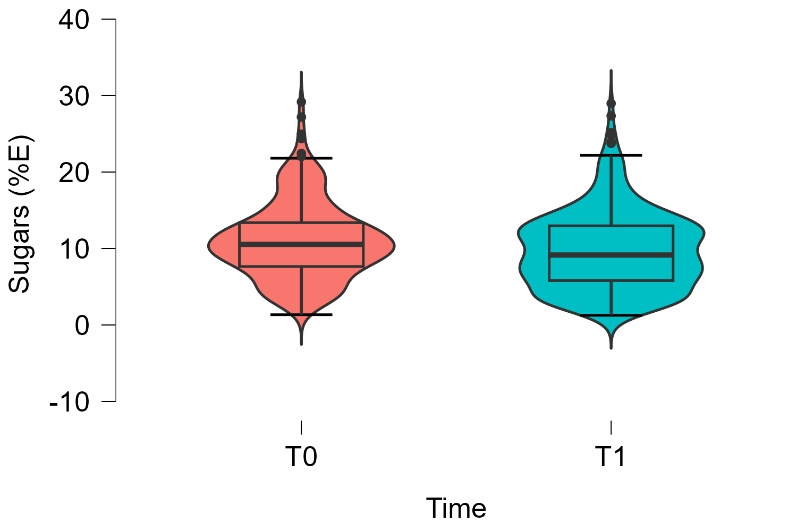

g) **SFA (%E)**

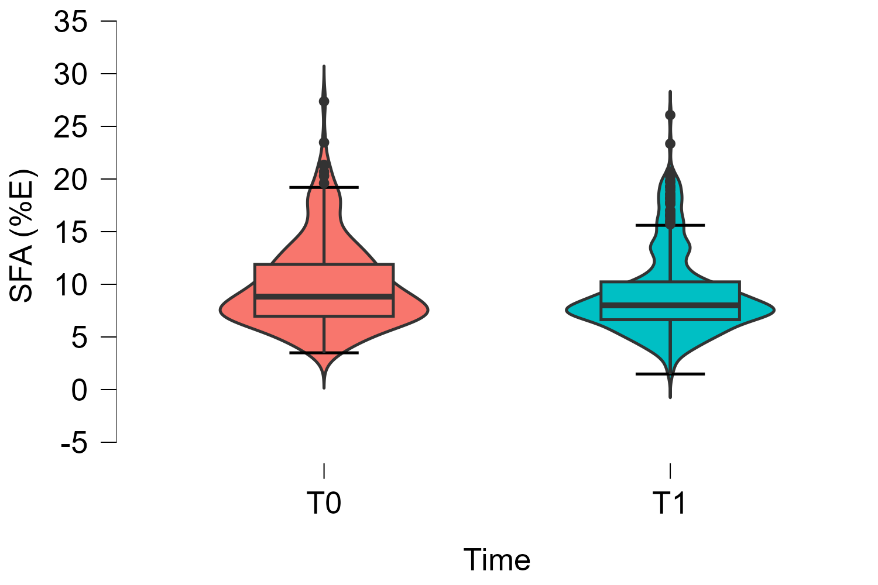

h) **MUFA (%E)**

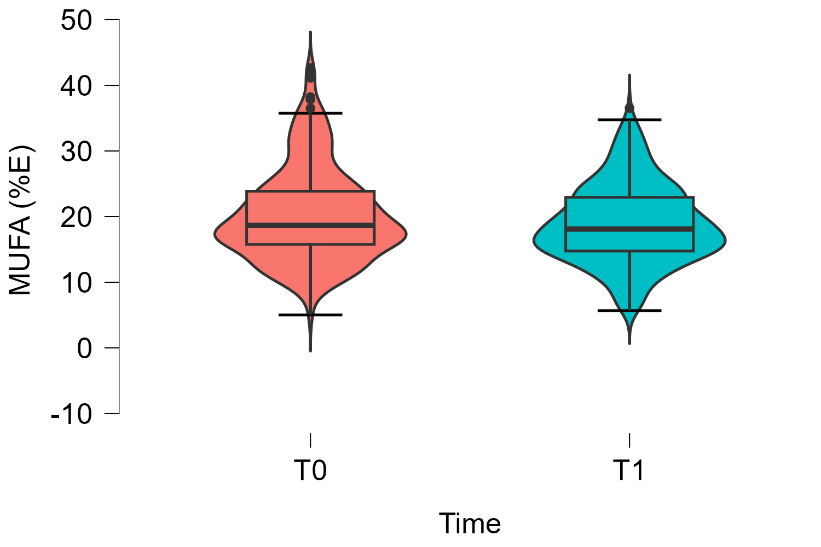

i) **PUFA (%E)**

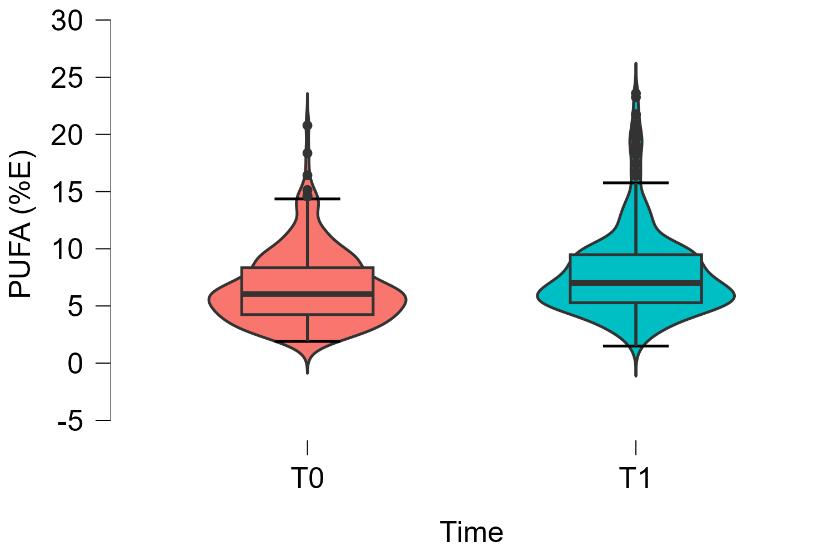

j) **EPA + DHA (mg)**

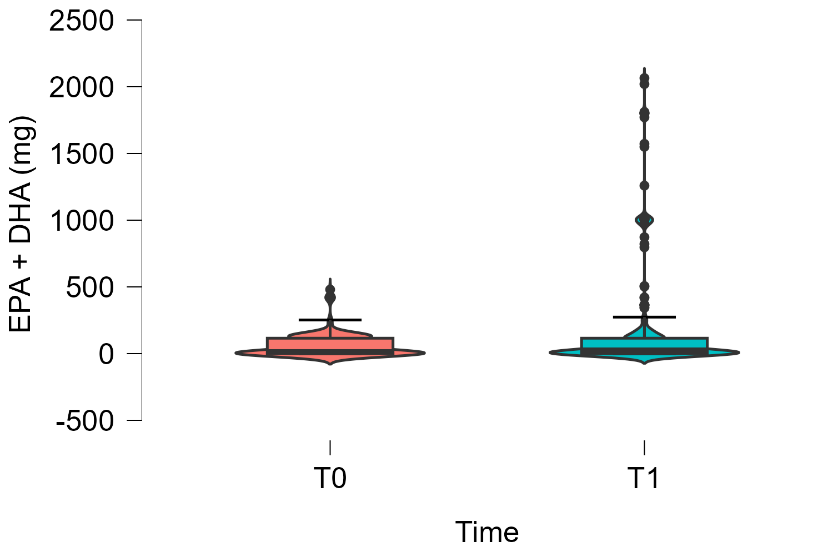

k) **Protein (g)**

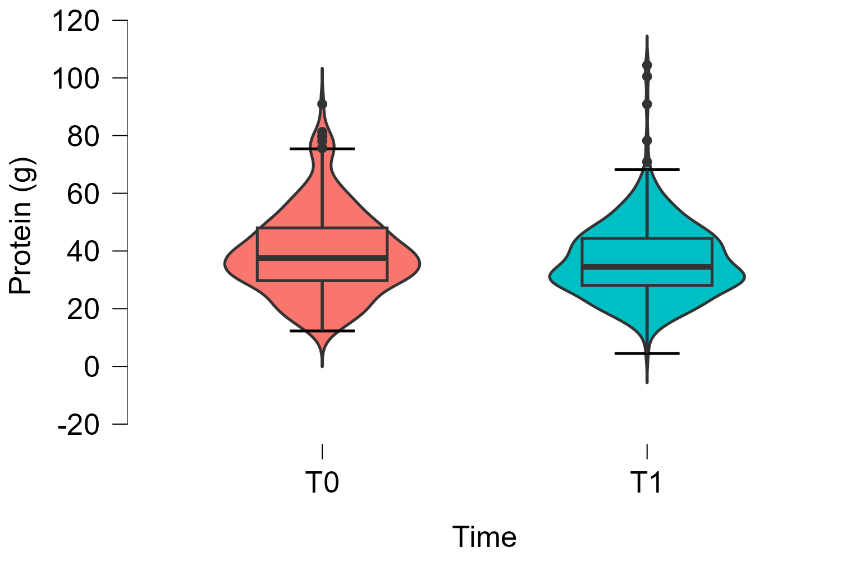

l) **Lipids (g)**

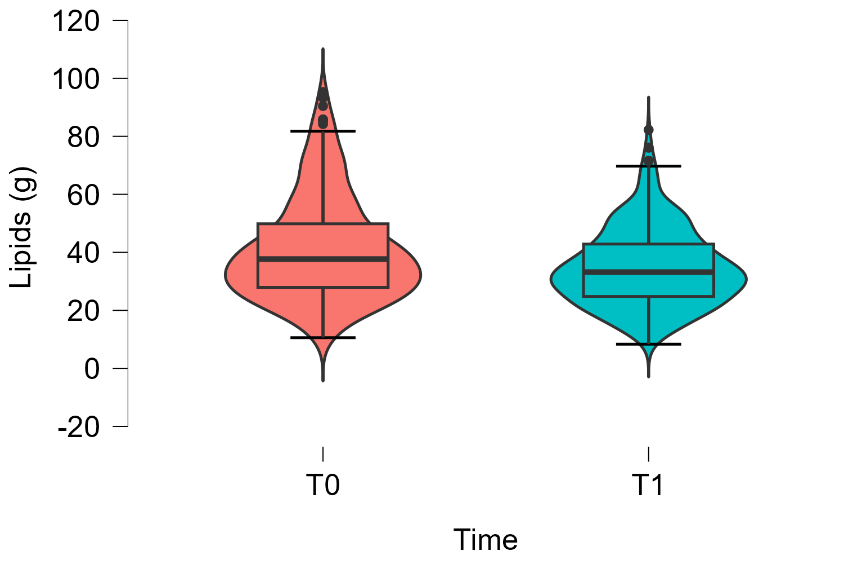

m) **Carbohydrates (g)**

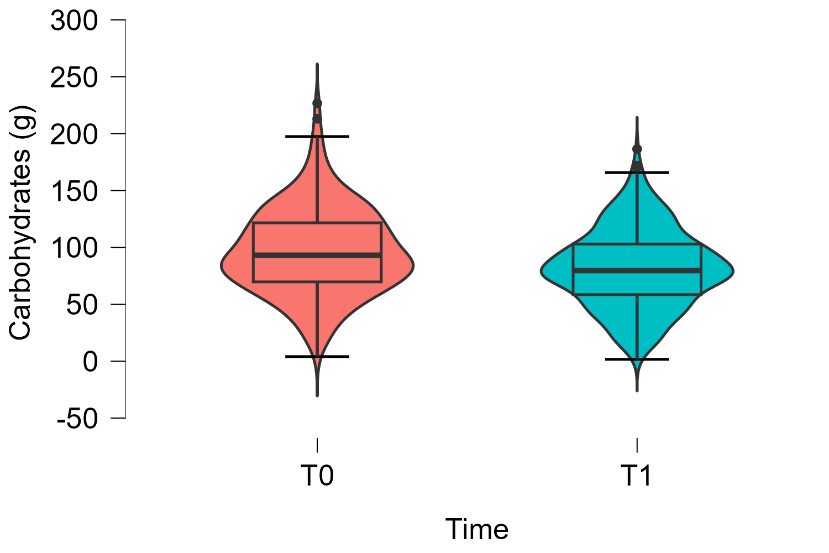

n) **Sugars (g)**

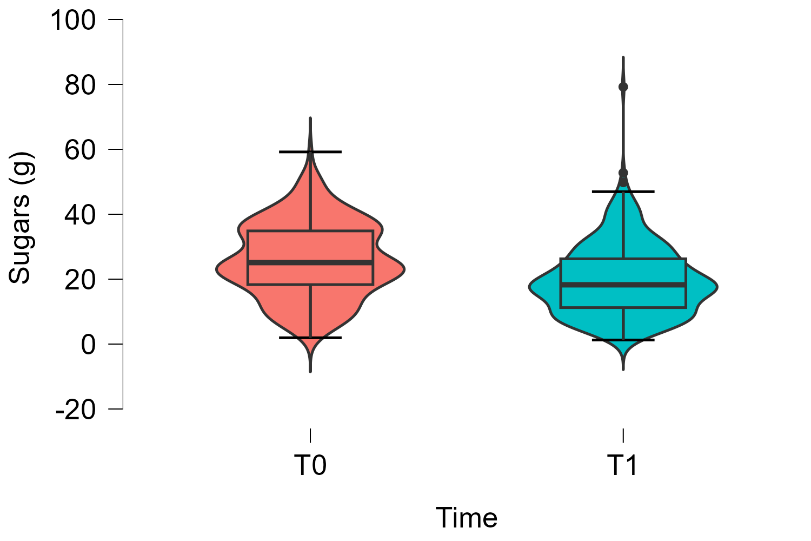

o) **Fibre (g)**

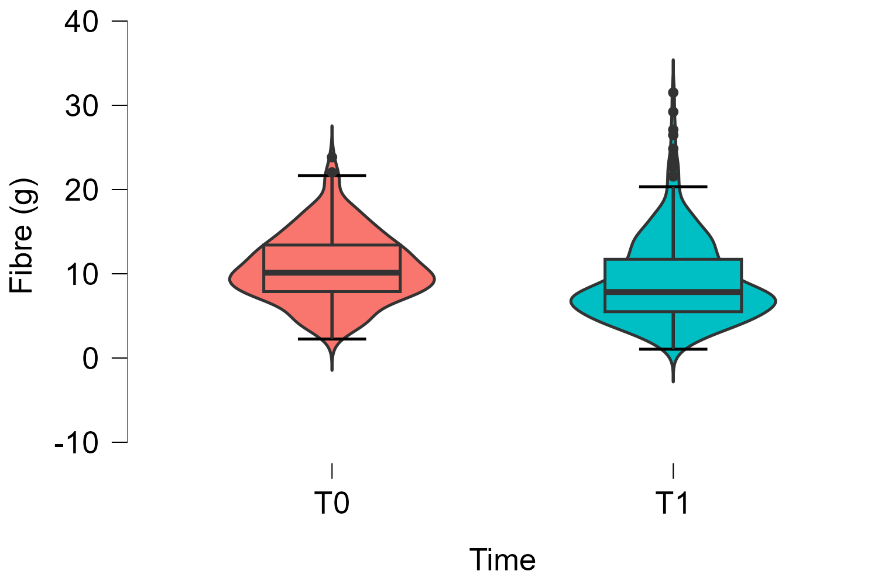

p) **SFA (g)**

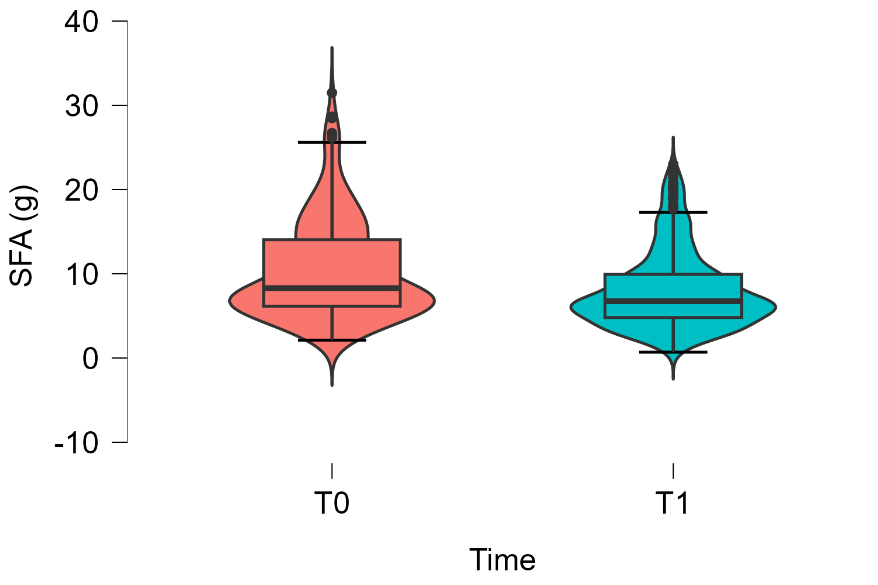

q) **MUFA (g)**

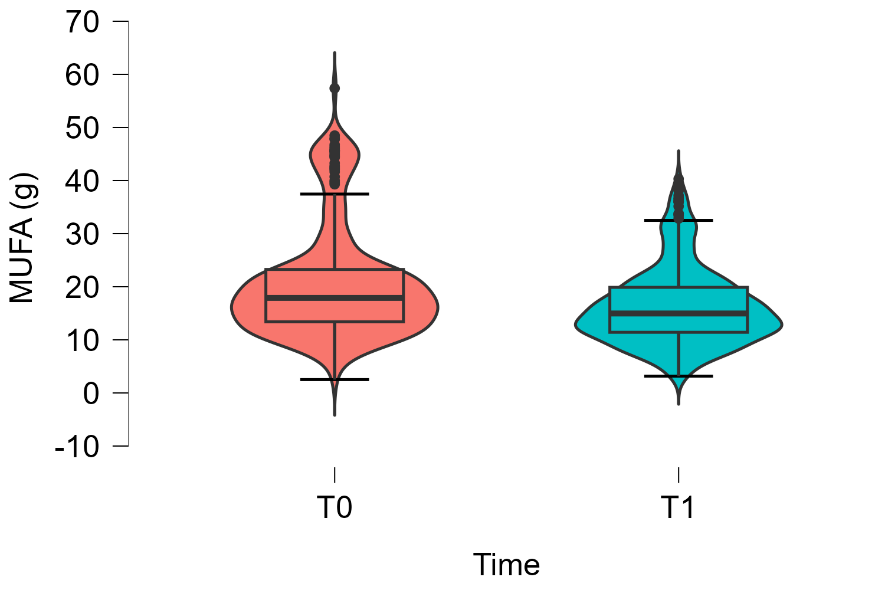

r) **PUFA (g)**

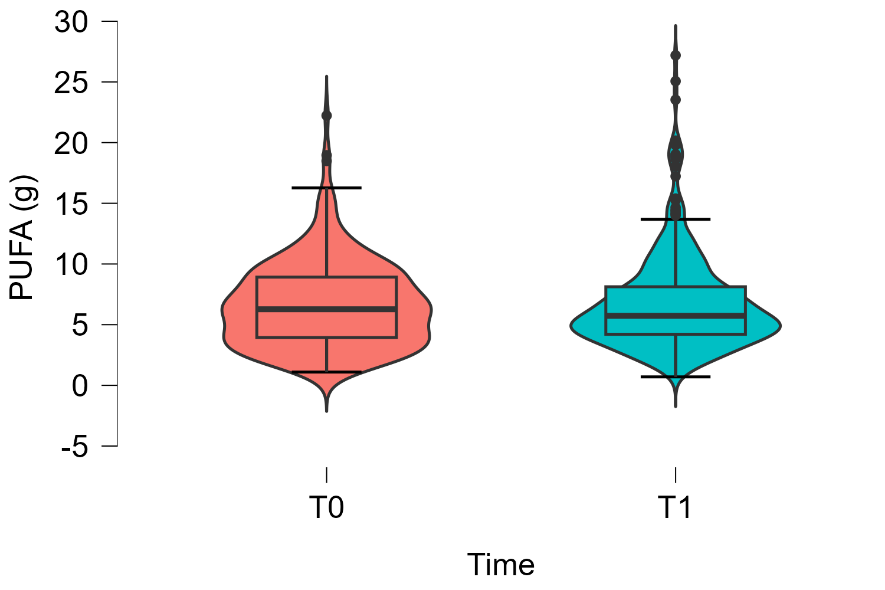

s) **CF (g CO2eq./tray)**

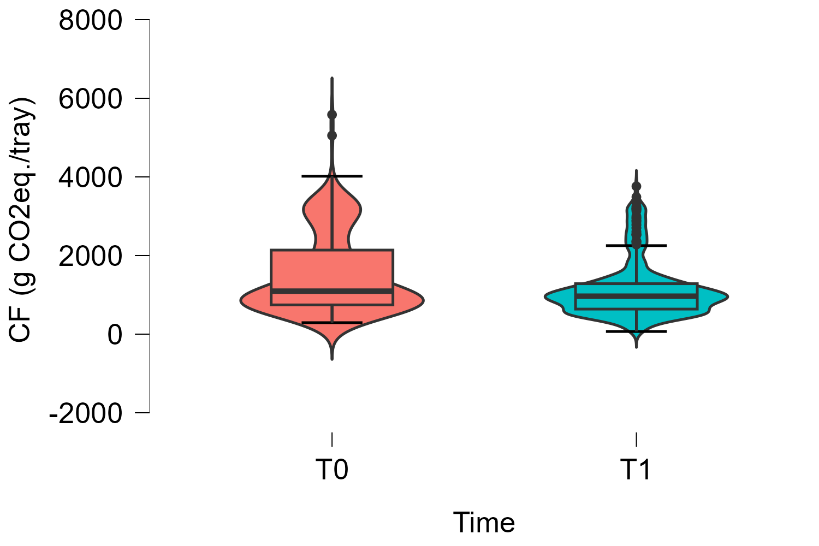

t) **WF (L H2O/tray)**

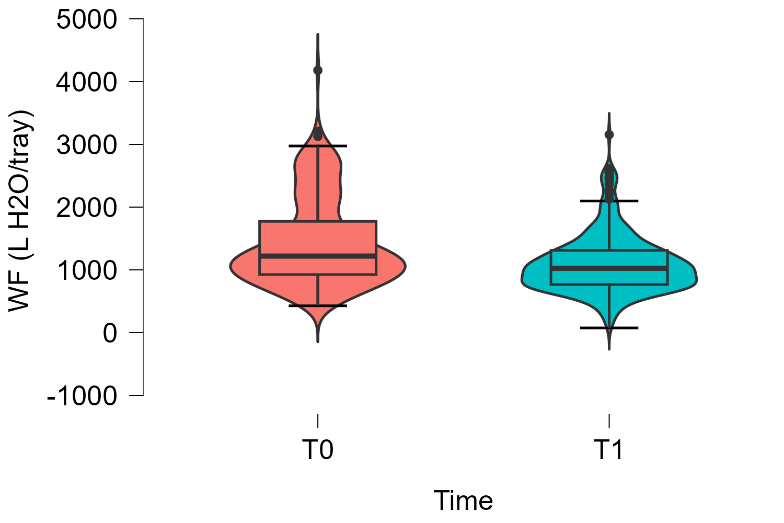

**Supplementary Figure 3. Distribution of nutritional and sustainability values in C3 at T0 and T1**

a) **Energy (kcal/tray)**

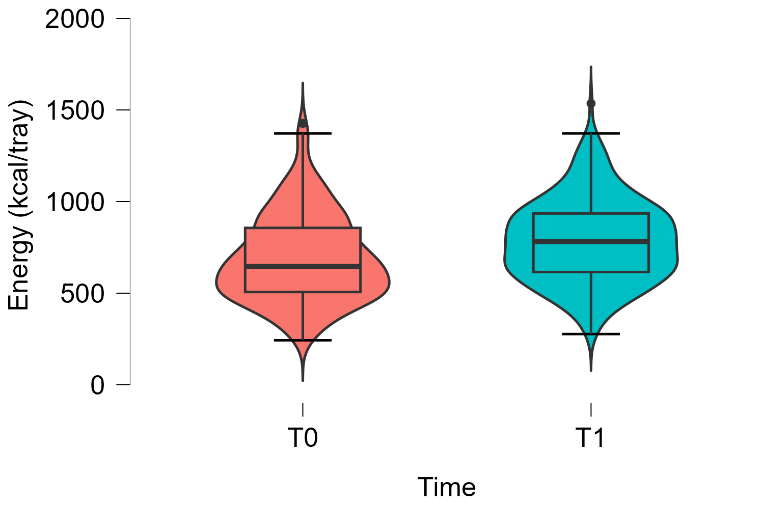

b) **Protein (%E)**

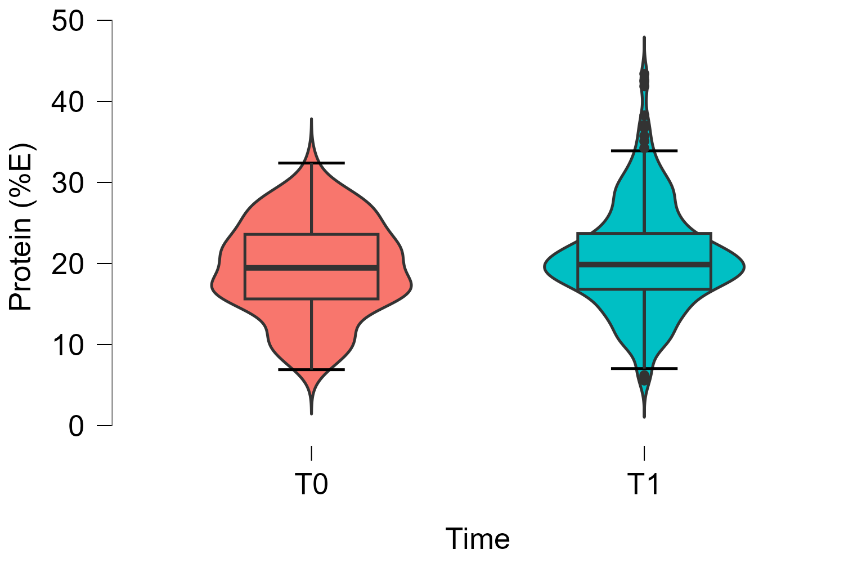

c) **Lipids (%E)**

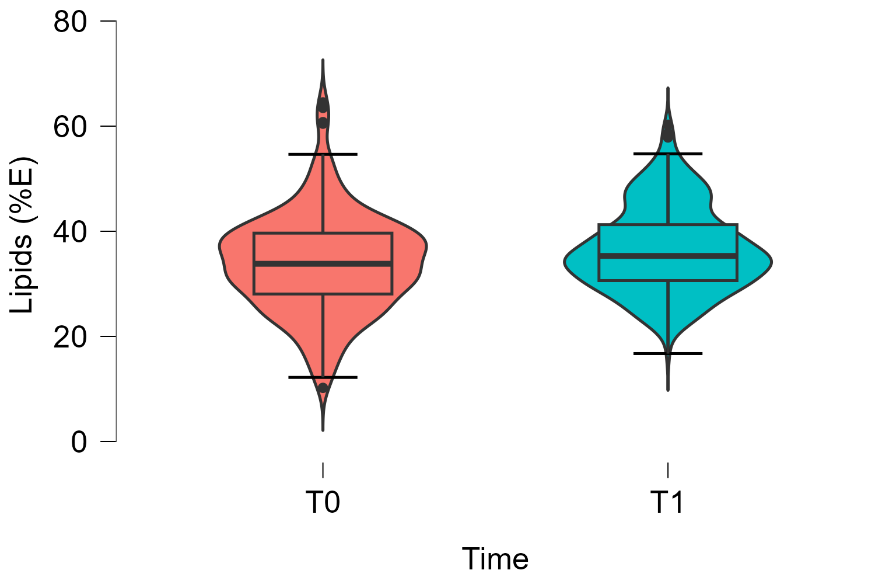

d) **Carbohydrates (%E)**

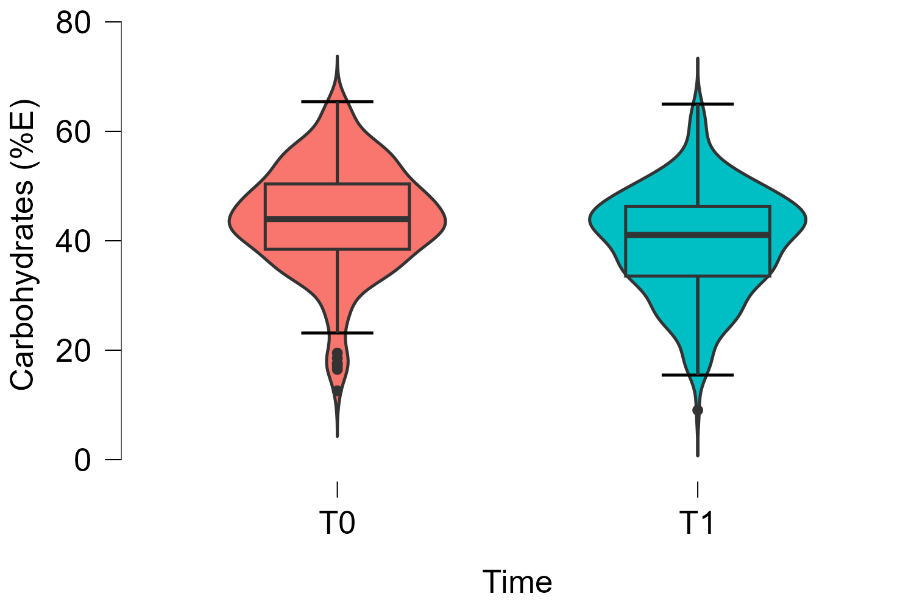

e) **Fibre (%E)**

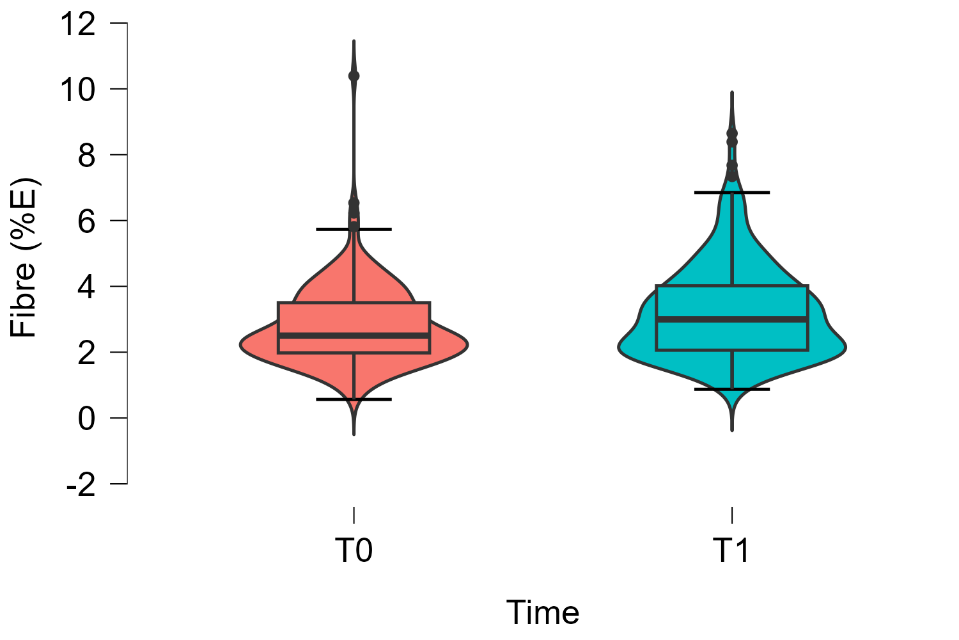

f) **Sugars (%E)**

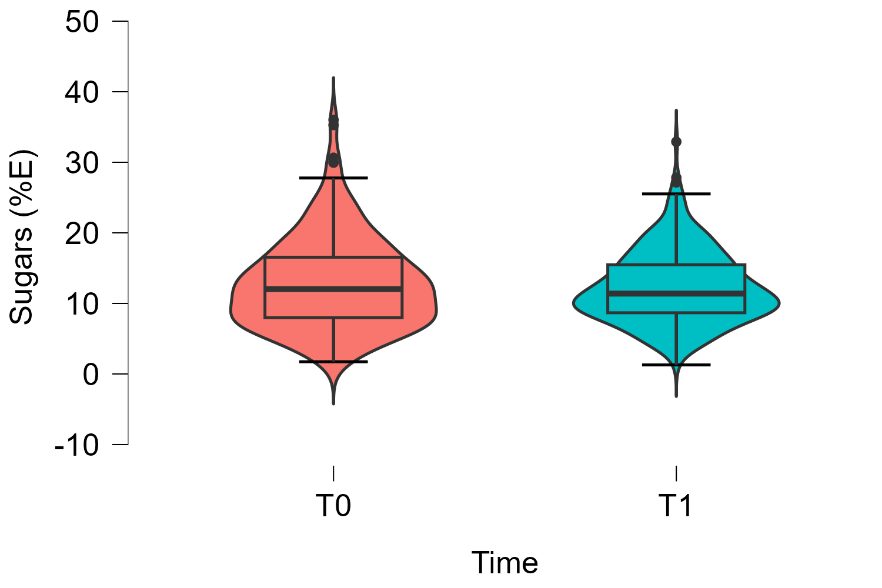

g) **SFA (%E)**

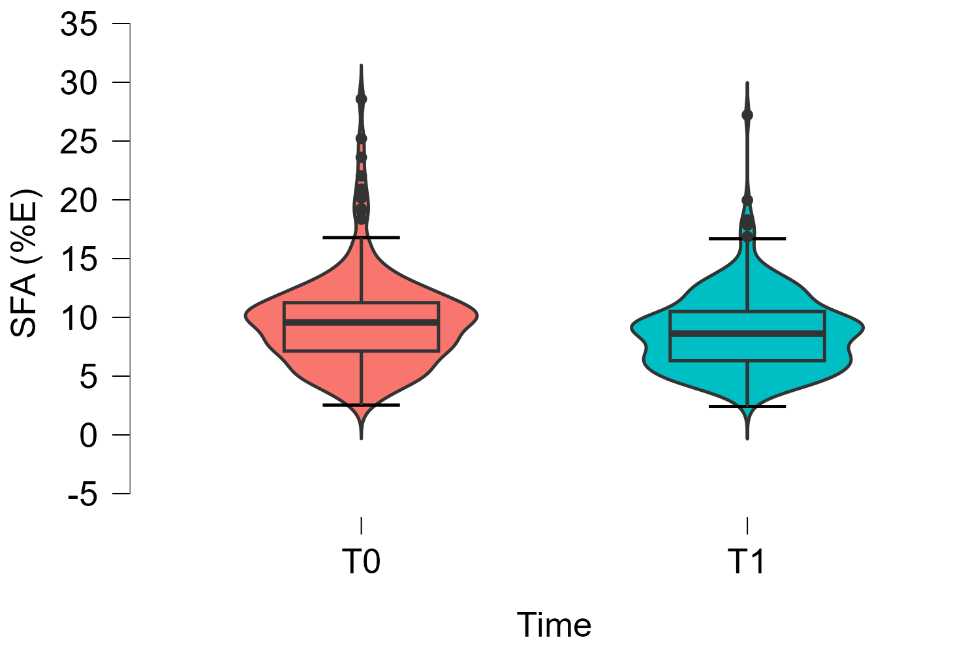

h) **MUFA (%E)**

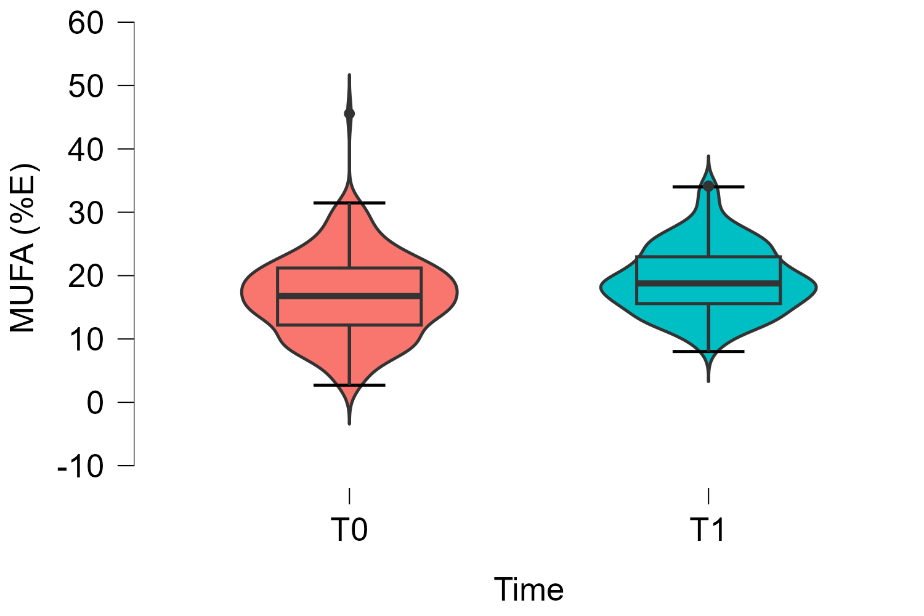

i) **PUFA (%E)**

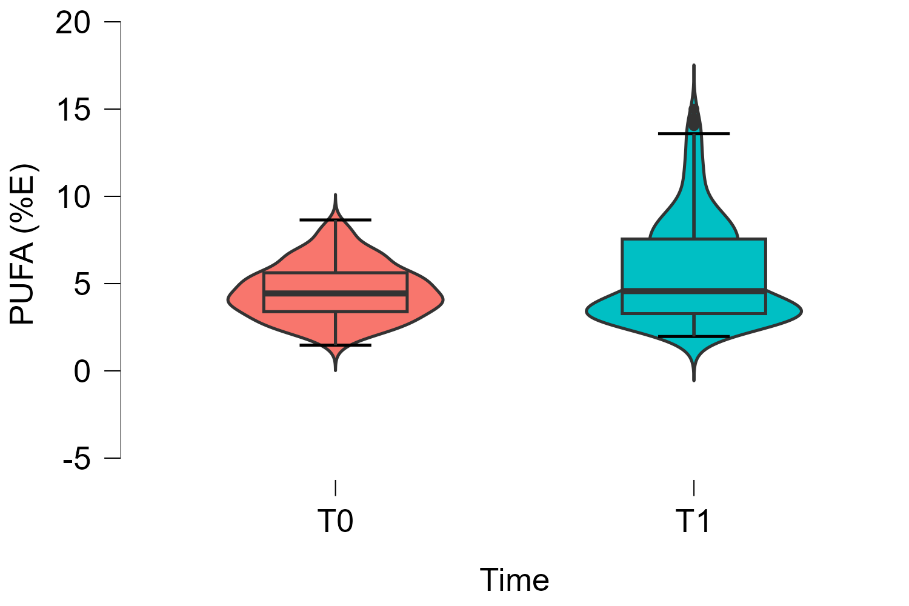

j) **EPA + DHA (mg)**

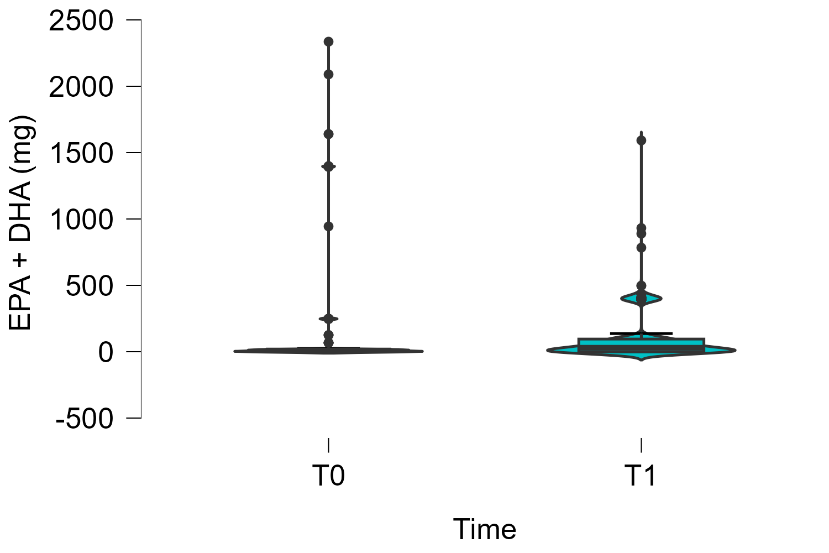

k) **Protein (g)**

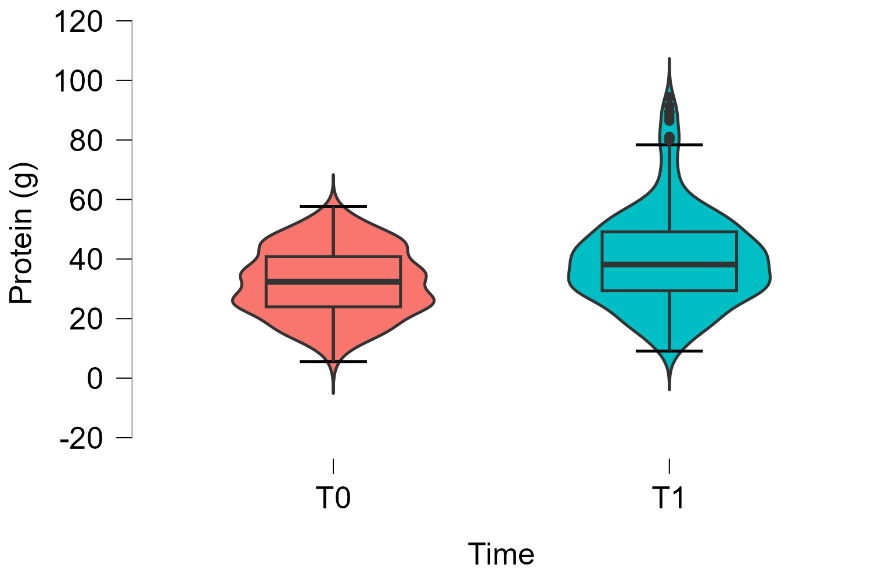

l) **Lipids (g)**

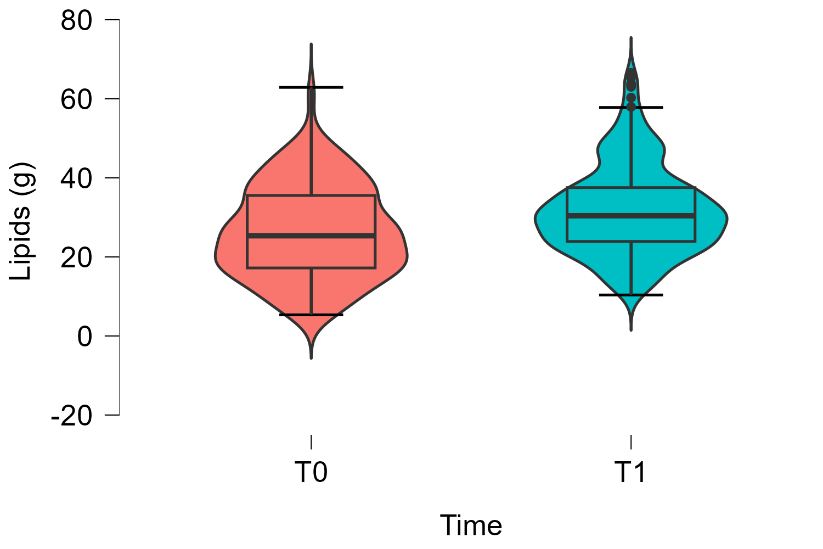

m) **Carbohydrates (g)**

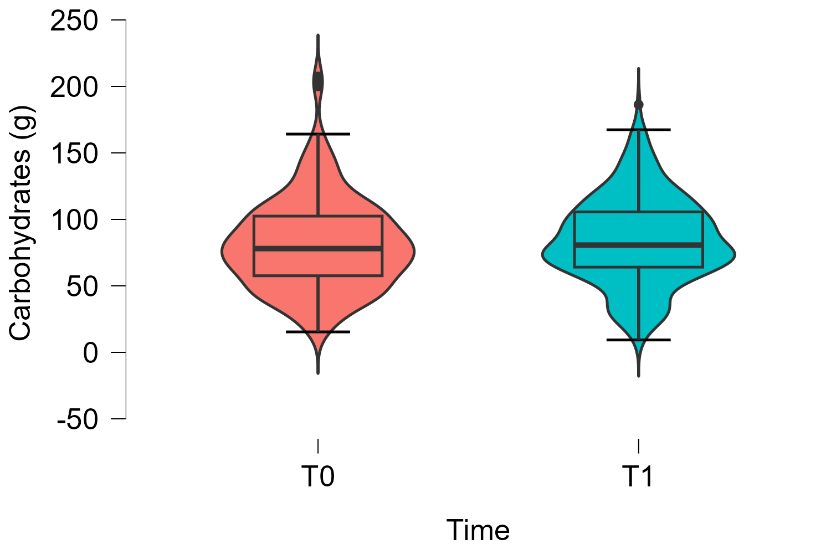

n) **Sugars (g)**

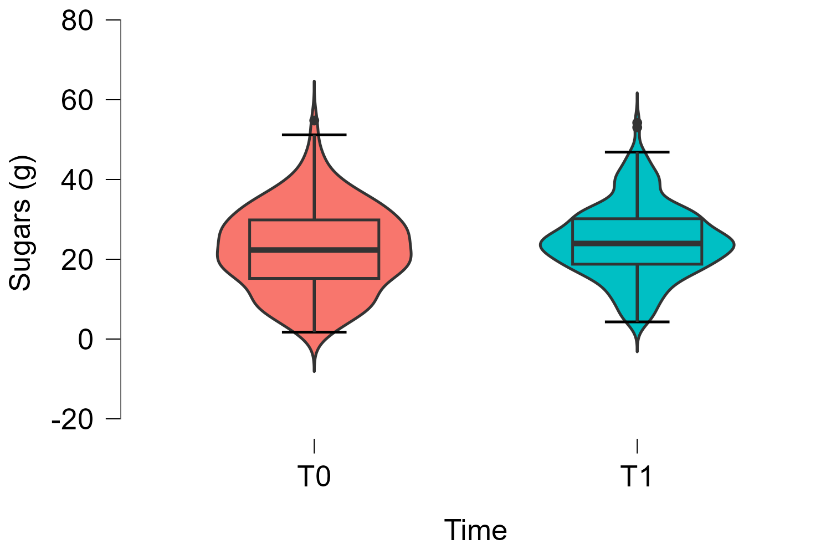

o) **Fibre (g)**

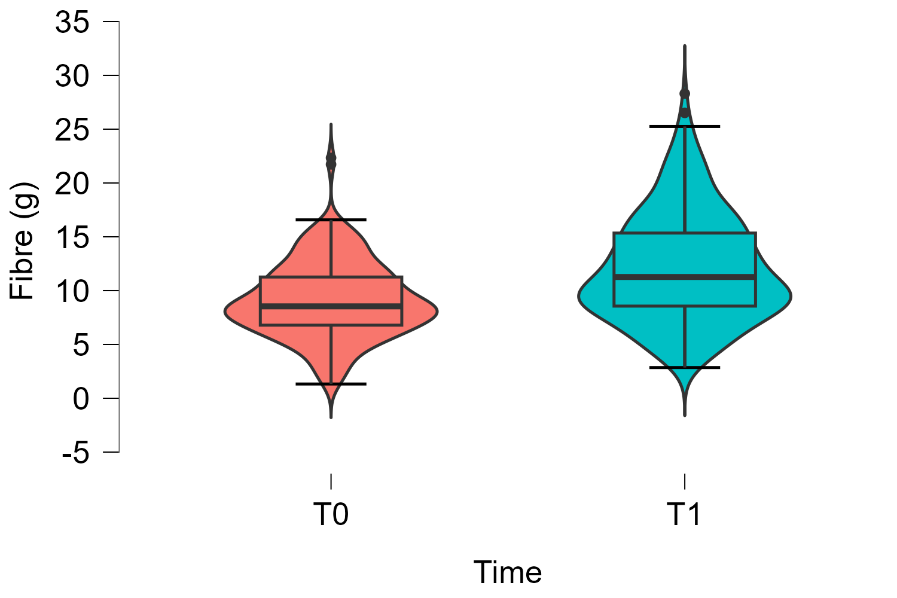

p) **SFA (g)**

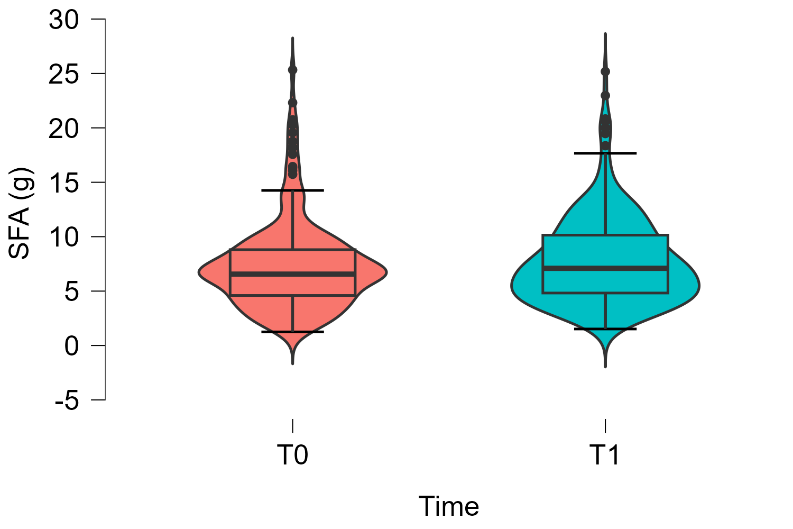

q) **MUFA (g)**

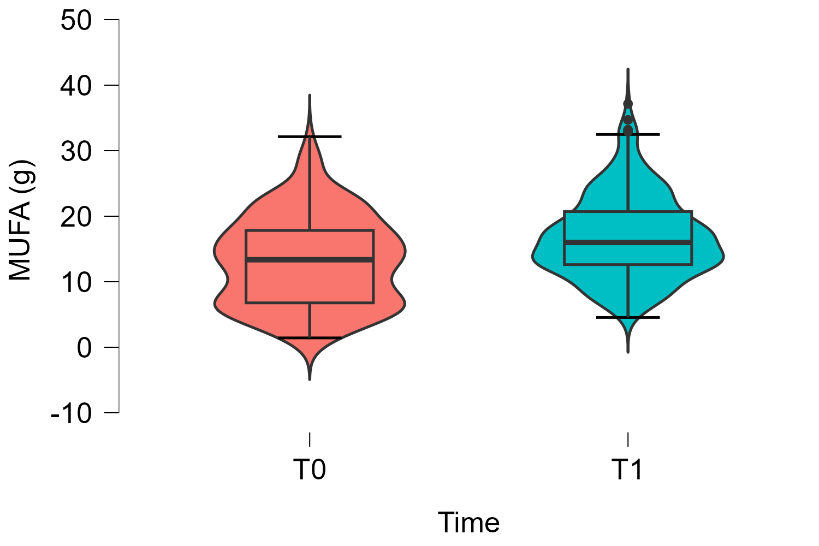

r) **PUFA (g)**

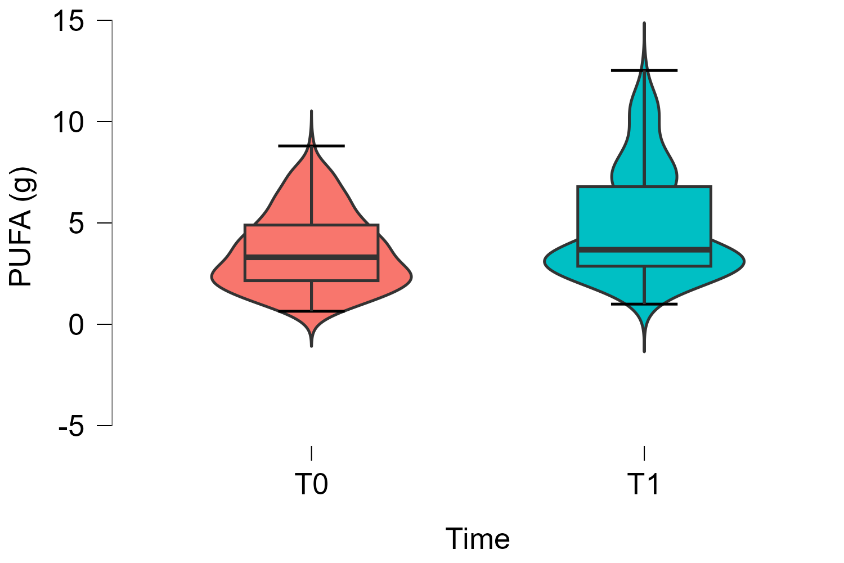

s) **CF (g CO2eq./tray)**

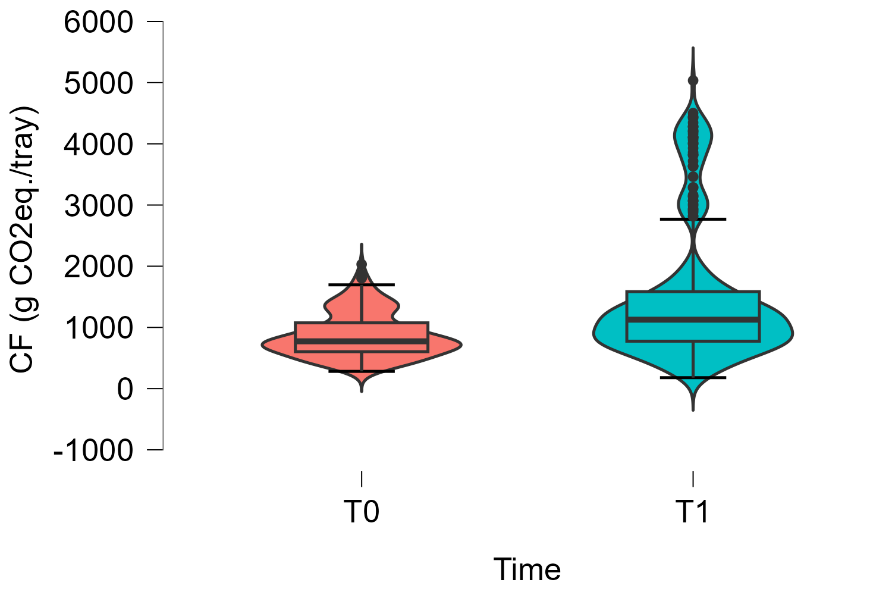

t) **WF (L H2O/tray)**

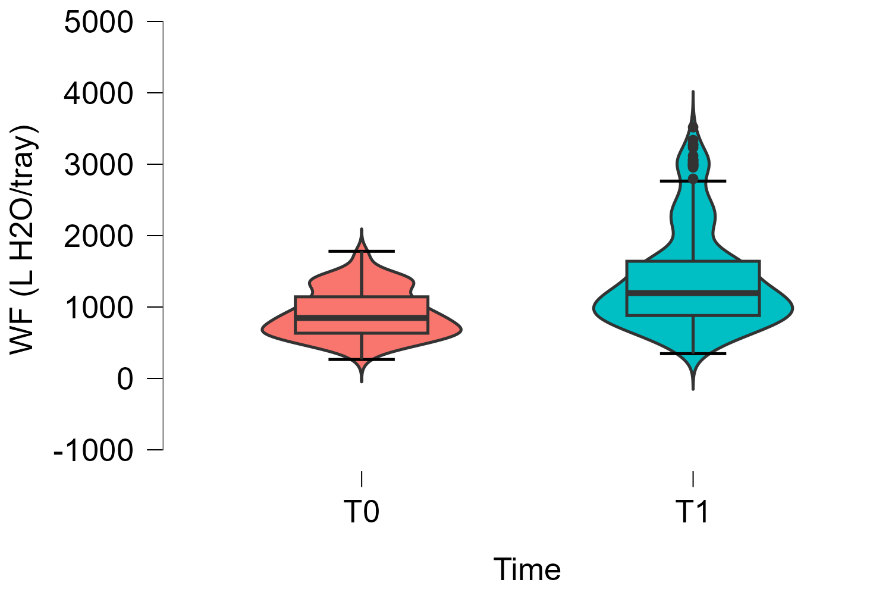
